# Working from home a few times a month to a few times a week is associated with higher psychological distress: between- and within-person analyses of a five-wave panel of employees in Japan

**DOI:** 10.64898/2026.10.21.26366098

**Authors:** Shun-ichiro Fuse, Hiroatsu Suga, Shuhei Ishikawa, Takahiro Tabuchi

## Abstract

**Purpose:** To estimate the association between the frequency of working from home (WFH) and psychological distress among employees in Japan, separating between-person from within-person differences, and to examine this association by sex and household circumstances.

**Methods:** We used five annual waves (2021–2025) of the nationwide Japan COVID-19 and Society Internet Survey (JACSIS), including 39,773 employees aged ≥18 years who worked in the past month (78,498 person-waves). The exposure was the frequency of WFH in the past month (seven categories). The outcomes were the Kessler 6 (K6) score (0–24), K6 ≥5, and K6 ≥13. We used within–between linear regression, overall and by sex and household circumstances at the first wave.

**Results:** Compared with no WFH, WFH 2–3 times a month, once a week, and 2–3 times a week was associated with K6 scores higher by 1.99, 1.80, and 0.67 points between persons and by 0.53 (95% CI 0.28, 0.77), 0.45 (0.20, 0.69), and 0.35 (0.10, 0.60) points within persons, and with a 3.5–4.6 percentage point higher probability of K6 ≥5 within persons. These within-person K6 differences were positive in all sex and household groups (0.20–0.63 points) but imprecise. For the other frequencies, the within-person differences were 0.03–0.20 points.

**Conclusion:** WFH 2–3 times a month or once a week, and also 2–3 times a week, was associated with higher psychological distress, both between and within persons. Support for employees who sometimes work from home should consider the frequency of WFH together with their work and family situations.

## Introduction

Working from home (WFH) spread as an emergency response to the COVID-19 pandemic and has continued since then, mainly in a form in which employees work from home on part of the week and at the workplace on the other days (Barrero et al. 2023). In Japan, the proportion of employees who telework has fallen from its level during the pandemic but has remained higher than before the pandemic, and telework on 1–4 days a week is becoming established (Ministry of Land, Infrastructure, Transport and Tourism 2025, 2026). How many times a week or a month to work from home has become a routine decision for workers and employers, and the frequency that workers prefer can differ from the frequency that employers allow (Liang et al. 2023). However, systematic reviews have not reached a consistent conclusion on the association between WFH and mental health (Lunde et al. 2022; Hall et al. 2023; Parent-Lamarche et al. 2026). A systematic review suggested that working from home on only part of the week may be more favorable in terms of psychosocial risk factors (Antunes et al. 2023), whereas a study in the United States estimated that the spread of WFH worsened mental health (Emanuel et al. 2026). Evidence is needed not only on whether people work from home but also on how the frequency of WFH is associated with mental health.

Most studies that examined associations by WFH frequency have been cross-sectional, and their results are inconsistent. In the United States, WFH on 1–4 days a week was associated with a slightly lower risk of depression (Leong et al. 2025). In Germany, fewer health complaints were found only when the extent of WFH was small (Mergener et al. 2025). In Sweden, employees who worked predominantly from home reported poorer health (Rosander 2026). In Norway, among employees who worked from home when needed or desired, distress was higher with more days of WFH (Trevino Garcia and Christensen 2025). In Japan, a cross-sectional study showed that a mismatch between the preferred and the actual frequency of WFH was associated with psychological distress (Otsuka et al. 2021).

Whether employees can work from home depends on their occupation, education, income, and company size (International Labour Organization 2021). Differences in psychological distress between people who work from home and those who do not also reflect these differences in background. When the same people are observed repeatedly, differences between people (between-person differences) can be examined separately from differences within the same person over time (within-person differences), and within-person estimates remove confounding by personal characteristics that do not change over time (Bell et al. 2019). In studies that followed the same people, WFH has been measured as full-time or part-time WFH (Gueguen and Senik 2023), the proportion of working hours spent at home (Botha et al. 2026), the extent of WFH over several months (Kornadt et al. 2025), or the extent to which an occupation allows WFH (Beaufils and Chung 2026; Emanuel et al. 2026). These measures do not correspond directly to the frequency that workers and employers decide on, that is, how many times a month or a week. Longitudinal studies in Japan followed participants for about 1 year between 2019 and 2021 and examined topics such as the start of WFH, occupational stress, and life satisfaction (Shimura et al. 2021; Sudo 2022; Ikegami et al. 2023). However, there is limited evidence on the within-person association between psychological distress and WFH at low frequencies, such as 2–3 times a month or once a week (Kageyama et al. 2026).

In addition, the association between WFH and mental health may differ by household circumstances and by sex. In the United States, the decline in mental health attributed to the spread of WFH was concentrated among people living alone (Emanuel et al. 2026). In a cross-sectional study in Japan, the association between WFH and psychological distress was stronger among people living with preschool children or with family members who needed care (Ikenouchi et al. 2023). In an Australian panel study, an association between WFH and better mental health among women was reported when 50–75% of working hours were spent at home, and this association was concentrated among women with poorer mental health (Botha et al. 2026). However, these findings are based on comparisons between occupations, cross-sectional data, or the proportion of working hours spent at home. Few studies have shown how much the between-person and within-person associations for each frequency of WFH differ by sex or household circumstances.

The aim of this study was to estimate the association between seven categories of WFH frequency and psychological distress among employees in Japan, separating between-person from within-person associations. We also examined differences in these associations by sex and household circumstances. We expressed the size of the associations as differences in the Kessler 6 (K6) score and as differences in the probability of scoring at or above the K6 screening thresholds.

## Methods

### Data

The Japan COVID-19 and Society Internet Survey (JACSIS) is an internet survey of residents across Japan. Participants are selected from the registered panel of an internet research company according to the distribution of sex, age, and region of residence, and the survey is conducted every year. Respondents to previous waves are surveyed again, and new respondents are added in each wave (Someko et al. 2026). We used five annual waves from 2021 to 2025, which were conducted between September 2021 and February 2026. In each wave, invalid responses, such as incorrect answers to attention-check items and inconsistent answers, were excluded (Someko et al. 2026). This study is reported in accordance with the STROBE statement.

### Participants

We included responses in each wave (hereafter, person-waves) from participants aged ≥18 years whose main job was as an employee and who actually worked in the past month. Employees were defined as regular staff or employees (including managers), dispatched workers, contract or entrusted employees, part-time or temporary workers, and students doing part-time work. Because of the response format of the survey, there were no missing values in the variables used in the analysis. The selection of participants is shown in Fig. S1, and the characteristics of participants by survey wave are shown in Table S1 (Supplementary Material).

## Measures

### Frequency of WFH

The frequency of WFH in the past month was measured in seven categories: none, once a month, 2–3 times a month, once a week, 2–3 times a week, 4–5 times a week, and almost daily. “None” (hereafter, no WFH) was used as the reference category. Telework at places other than home was not included.

### Psychological distress

Psychological distress in the past 30 days was measured with the Japanese version of the K6 (Kessler et al. 2002; Furukawa et al. 2008). The main outcome was the total score of the six items (range 0–24; higher scores indicate more distress). As secondary outcomes, we used whether the score met K6 ≥5 (Prochaska et al. 2012) and K6 ≥13 (Kessler et al. 2003).

### Covariates

Sex, age, region of residence, and education were fixed at the values from the person’s first response in the 2021–2025 surveys. Age was converted to the 2021 equivalent and grouped into seven categories, region into eight regions, and education into four categories. The following covariates were used as values at each wave: occupation (10 categories), industry (19 categories), employment type (regular, dispatched, contract or entrusted, and part-time or temporary), company size (5 categories), household type (living alone; living with a spouse or partner and no child <18 years; living with a spouse or partner and a child <18 years; and other), annual household income (8 categories), and survey wave.

The definitions of the categories are given in the Supplementary Methods (Supplementary Material).

## Statistical analysis

### Main analysis

To estimate the between-person and within-person associations simultaneously, we used a within–between model (Curran and Bauer 2011; Bell et al. 2019). Let *i* denote the person and *t* the survey wave, and let *D_kit_* (*k* = 1, … ,6) be indicator variables for the six frequencies of WFH, with no WFH as the reference. **C***_it_* is a vector of time-varying covariates and survey wave indicators, and **Z***_i_* is a vector of characteristics fixed at the first response. A bar denotes the mean for each person over the waves in which the person was in the analysis.

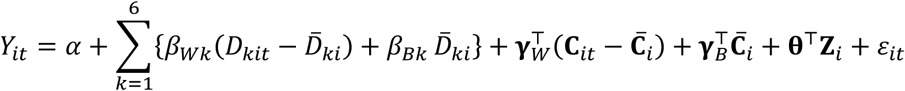

*β_Wk_* (the within-person association) represents the difference in K6 within the same person between WFH at frequency *k* and no WFH. It is estimated from all changes in the frequency category within persons, including changes between two frequencies of WFH (for example, from once a week to 2–3 times a week); therefore, persons who worked from home in all of their waves also contribute if their frequency changed. Because the comparison is made within the same person, time-invariant differences specific to each person are removed (Bell et al. 2019; Wooldridge 2019). *β_Bk_* (the between-person association) represents the difference in K6 between people who worked from home at frequency *k* in all of their wave in the analysis and people with no WFH in all of their waves, with other characteristics held constant. The model was estimated by ordinary least squares, and standard errors were clustered by person. For K6 ≥5 and K6 ≥13, we used linear probability models of the same form and expressed the differences in probability in percentage points. Estimates are presented with 95% confidence intervals (CIs).

### Differences by sex and household circumstances

As an additional analysis, we estimated the between-person and within-person associations with the K6 score separately by sex and by household circumstances at the first wave. Household circumstances were classified into three groups based on the response at each person’s first wave in the analysis: living with others and no child <18 years, living alone, and living with a child <18 years. The same group was assigned to all of that person’s waves. We fitted the same model as in the main analysis to each group and estimated all coefficients separately by group (in the analysis by sex, sex was removed from the covariates).

### Supplementary analyses

We estimated the association between the frequency of WFH at the current wave and K6 at the next survey wave, using as the unit of analysis the same person’s responses in two adjacent surveys (hereafter, a pair) and adjusting for K6 and covariates at the current wave (Supplementary Methods). We also repeated the main analysis under four conditions: (1) adjustment only for sex, age, region, education, and survey wave; (2) restriction to person-waves working ≥30 hours a week; (3) restriction to person-waves whose main work was desk work; and (4) weighting by the inverse of the number of waves, so that the weights sum to the same total for each person.

Analyses were conducted with R version 4.5.3 and the fixest package version 0.14.1. Generative artificial intelligence tools (Claude, Anthropic) were used to assist in writing part of the analysis code and editing the manuscript. The authors checked all code, results, and text and take full responsibility for the content.

## Results

### Participant characteristics

The analytic sample comprised 78,498 person-waves from 39,773 persons (Fig. S1). Of these persons, 19,056 (47.9%) were observed in two or more waves, and 4,879 of them (25.6%) changed their WFH frequency category at least once during the observation period. The frequency of WFH in each survey wave and the transitions between adjacent waves are shown in Fig. S2. WFH in the past month was reported in 21.4% of person-waves, and this proportion fell from 24.2% in 2021 to 19.4–20.6% in 2023–2025 (Table S1).

Compared with those who did not work from home, those who worked from home were more likely to be men, to have a university degree or higher, to be regular employees, to work for a company with ≥1,000 employees, and to do mainly desk work (Table 1). They were also more likely to live in the Kanto region, which includes Tokyo (Table S2). At the first wave in the analysis, the mean K6 score was 4.9 for no WFH, 6.3 for WFH 2–3 times a month, and 6.4 for WFH once a week.

**Table 1.**
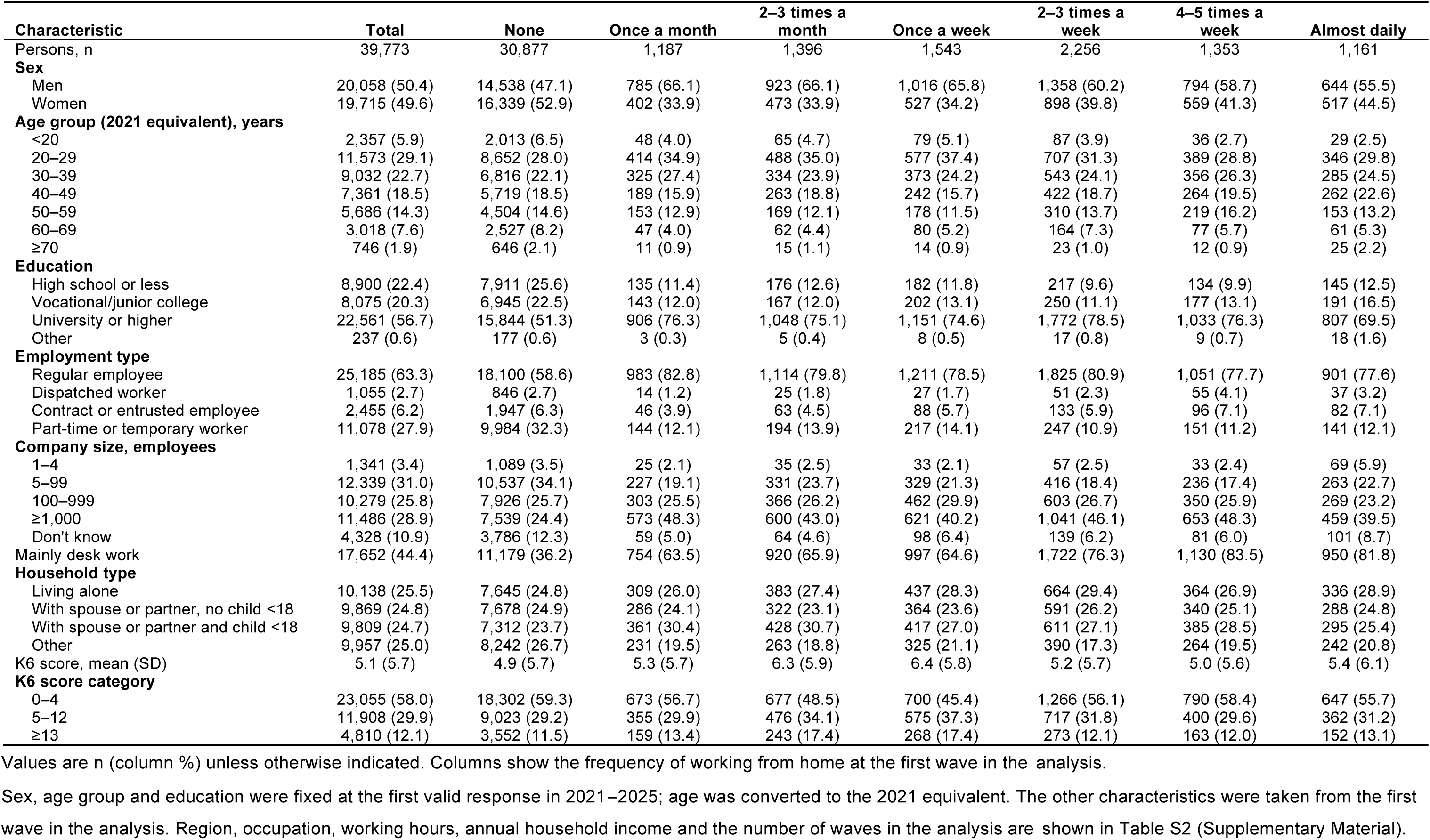
Characteristics of participants at their first wave in the analysis, by frequency of working from home.

### Frequency of WFH and psychological distress: between-person and within-person associations

Between persons, compared with no WFH, WFH 2–3 times a month, once a week, and 2–3 times a week was associated with K6 scores that were higher by 1.99 points (95% CI 1.61, 2.37), 1.80 points (1.45, 2.14), and 0.67 points (0.37, 0.96), respectively. For the other frequencies, the differences were 0.31–0.73 points (Fig. 1).

**Fig. 1.**
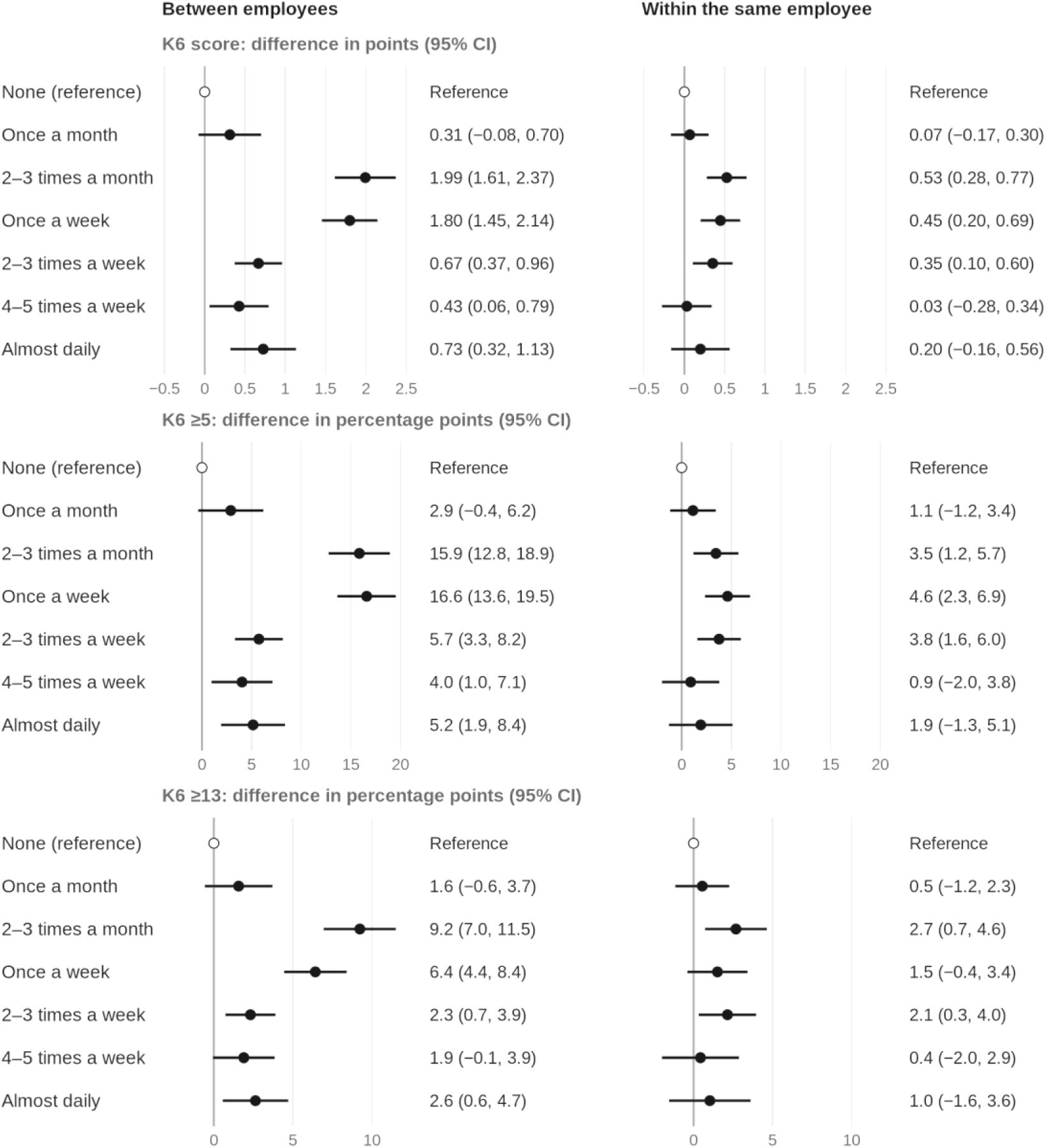
Frequency of working from home and psychological distress: between-person and within-person associations. For each frequency, the difference compared with no working from home is shown for the K6 score (top), the probability of K6 ≥5 (middle), and the probability of K6 ≥13 (bottom), as point estimates with 95% confidence intervals. Values on the right are point estimates (95% confidence intervals)

Within persons, compared with no WFH, WFH 2–3 times a month, once a week, and 2–3 times a week was associated with K6 scores that were higher by 0.53 points (0.28, 0.77), 0.45 points (0.20, 0.69), and 0.35 points (0.10, 0.60), respectively. The differences for once a month, 4–5 times a week, and almost daily were 0.03–0.20 points, and the 95% CI for almost daily (0.20 points) ranged from −0.16 to 0.56 points.

The outcomes based on thresholds showed the same pattern. Within persons, WFH from 2–3 times a month to 2–3 times a week was associated with a 3.5–4.6 percentage point higher probability of K6 ≥5. The differences in the probability of K6 ≥13 were 2.7 percentage points (0.7, 4.6) for 2–3 times a month, 1.5 percentage points (−0.4, 3.4) for once a week, and 2.1 percentage points (0.3, 4.0) for 2–3 times a week. All coefficients are shown in Table S3.

### Differences by sex and household circumstances

There were 20,058 men and 19,715 women. By household circumstances at the first wave, 19,202 persons were living with others and had no child <18 years, 10,138 were living alone, and 10,433 were living with a child <18 years. The proportion of person-waves with WFH was 25.9% among men, 16.3% among women, and 18.6–24.5% across the household groups. Among persons observed in two or more waves, the proportion who changed their WFH frequency category was 19.6–30.8% across the groups (Table S6).

The within-person point estimates for WFH from 2–3 times a month to 2–3 times a week were positive in both sexes and in all three household groups (0.20–0.63 points; Fig. 2). Among men, the estimates were 0.63 points (0.34, 0.92) for 2–3 times a month, 0.45 points (0.15, 0.76) for once a week, and 0.25 points (−0.06, 0.56) for 2–3 times a week; among women, they were 0.25 points (−0.22, 0.71), 0.44 points (0.02, 0.86), and 0.55 points (0.14, 0.96), respectively. By household circumstances, the estimates were 0.45–0.59 points for those living with others and no child <18 years, 0.20–0.51 points for those living alone, and 0.33–0.45 points for those living with a child <18 years. The widths of the 95% CIs of the group-specific estimates were 0.59–0.99 points, indicating more uncertainty than in the main analysis (0.49–0.50 points).

**Fig. 2.**
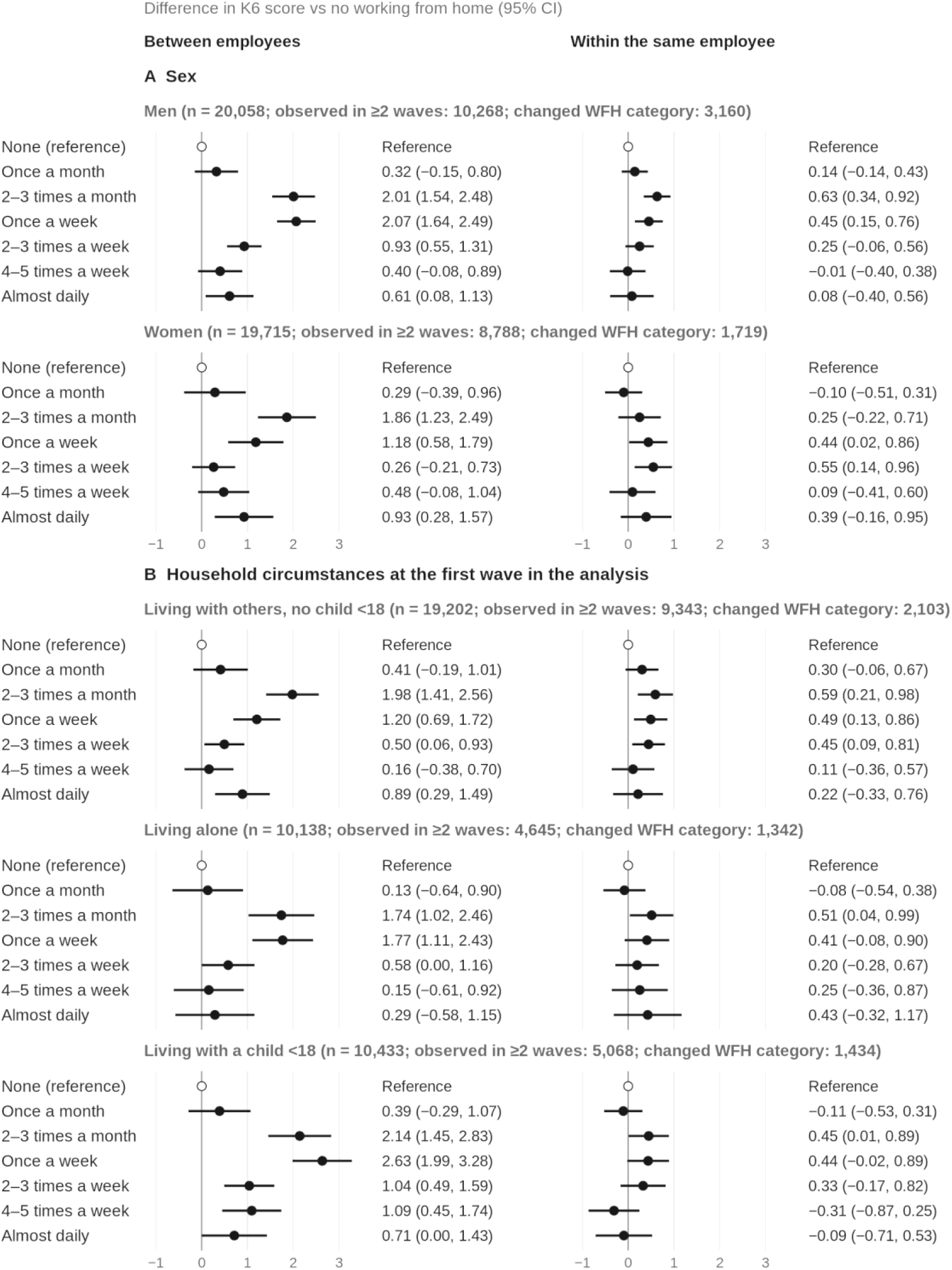
Between-person and within-person associations of the frequency of working from home with the K6 score, by sex (A) and by household circumstances at the first wave in the analysis (B). Differences in the K6 score compared with no working from home are shown as point estimates with 95% confidence intervals. Values on the right are point estimates (95% confidence intervals). The left column shows between-person associations and the right column shows within-person associations; the horizontal axis is the same in all panels. Household circumstances were classified by the response at each person’s first wave in the analysis, and the same group was assigned to all of that person’s waves. Within each group, the models were adjusted for the same covariates as in the main analysis (except sex in the analysis by sex), and all coefficients were estimated separately by group. Standard errors were clustered by person

Between persons, WFH 2–3 times a month and once a week was associated with higher K6 scores than no WFH in all groups (1.18–2.63 points). The between-person differences for 2–3 times a week were 0.26–1.04 points (Fig. 2).

### Supplementary analyses

Of the 61,936 person-waves in the analytic sample in 2021–2024, 37,043 (59.8%; 18,646 persons) had K6 observed at the next survey wave and formed pairs (Tables S4 and S5). After adjustment for K6 and covariates at the current wave, the differences in K6 at the next survey wave compared with no WFH were −0.01 to 0.22 points. Without adjustment for K6 at the current wave, the differences were 1.05 points (0.68, 1.41) for 2–3 times a month and 0.85 points (0.51, 1.19) for once a week (Fig. S3).

When the models were adjusted only for basic characteristics and when they were weighted by the inverse of the number of waves, the within-person estimates for 2–3 times a month, once a week, and 2–3 times a week were close to those in the main analysis (0.55, 0.46, and 0.37 points; and 0.54, 0.42, and 0.40 points, respectively). When the analysis was restricted to person-waves working ≥30 hours a week, the estimates were 0.44, 0.37, and 0.25 points, and when it was restricted to person-waves with mainly desk work, they were 0.37, 0.32, and 0.17 points; these estimates were smaller than those in the main analysis (Fig. S4).

## Discussion

### Main findings

In this five-wave panel of employees in Japan, WFH at low frequencies (2–3 times a month or once a week) was associated with higher psychological distress than no WFH. This was also the case for WFH 2–3 times a week. The association was seen not only in comparisons between people but also in comparisons of the same person at different times. The within-person differences were 0.35–0.53 points in the K6 score and 3.5–4.6 percentage points in the probability of K6 ≥5. In the analyses by sex and by household circumstances at the first wave, the within-person point estimates for these three frequencies were consistently positive. These results suggest the need to consider not only the characteristics of people who work from home but also the periods in which the same person’s frequency of WFH changes.

### Comparison with previous studies

The results of cross-sectional studies by WFH frequency have been inconsistent. In the United States, WFH on 1–4 days a week was associated with a slightly lower risk of depression (Leong et al. 2025), which is the opposite direction to our findings. In Norway, employees who worked from home when needed or desired had lower distress than those who did not work from home, whereas among these employees, distress was higher with more days of WFH (Trevino Garcia and Christensen 2025). This comparison differs from ours, in which each frequency was compared with no WFH. To understand these results, it is important to consider which working arrangements are being compared, as well as the WFH policies and the reasons for using WFH.

Among longitudinal studies, no clear within-person association was reported for the extent of WFH over several months in five European countries (Kornadt et al. 2025), and in Switzerland, the transition to WFH was not associated with health indicators (Altwicker-Hámori et al. 2026). By using detailed categories of the number of times per month or week, our study showed within-person associations for WFH from 2–3 times a month to 2–3 times a week. This adds evidence that corresponds to the frequencies of WFH that are chosen in everyday working life.

### Interpretation

Working from home on some days of the month or week gives employees an opportunity to balance the demands of work and family, but it also involves switching between work locations and managing the boundary between work and private life. A previous study reported that setting fixed times for WFH was associated with better mental health through better work–life balance (Jiang et al. 2022). It is also possible that employees use WFH in periods when their workload, physical condition, or family situation has changed. A mismatch between the preferred and the actual frequency of WFH has been associated with psychological distress (Otsuka et al. 2021) and insomnia (Hamada et al. 2024). The associations observed for WFH 2–3 times a month and once a week may also reflect the burden of switching between work locations and the circumstances that led employees to use WFH.

By household circumstances, the within-person point estimates for WFH from 2–3 times a month to 2–3 times a week were positive not only among those living alone and those living with a child <18 years but also among those living with others and no child <18 years (0.45–0.59 points). The point estimates for these three frequencies were in the same direction for men and women. Between persons, WFH 2–3 times a month and once a week was associated with higher K6 scores in all groups. Given these findings, attention to the work and life circumstances in which a person works from home on some days of the month or week, in addition to household composition and sex, may help to understand the associations.

The between-person differences (about 2 points for 2–3 times a month and once a week) were larger than the within-person differences (about 0.5 points). Presenting the two associations separately is useful because it captures both the possibility that differences in the backgrounds of people who use WFH contribute to the between-person association and the fact that the level of distress differs within the same person between periods with different frequencies of WFH (Bell et al. 2019).

### Implications

The association of WFH 2–3 times a month or once a week with higher psychological distress suggests that employees who work from home relatively infrequently should also be considered in workplace mental health support. In workplace consultations, assessing the reasons for WFH, workload, and work–family balance, rather than WFH frequency alone, may help identify employees’ needs and guide appropriate support.

## Strengths and limitations

A strength of this study is that we observed employees in Japan in five annual surveys from late 2021 to early 2026, estimated the between-person and within-person associations of seven categories of WFH frequency with psychological distress separately in one model, and examined these associations by sex and household circumstances within the same framework.

This study has three limitations. First, WFH and the K6 referred to the same past month, so their temporal order cannot be determined. Confounding by factors that change over time, such as workload, physical condition, and family circumstances, may remain even in within-person comparisons. Second, we did not measure the reasons for WFH, whether WFH days were fixed or flexible, employees’ preferences, or their discretion over WFH. Household circumstances were classified by the first response, and in each group, 14–19% of the persons observed in two or more waves moved to another group during follow-up (Table S6). We also did not distinguish whether there were preschool children or family members who needed care. In addition, sex and household circumstances are not direct measures of roles within the household or of social isolation, and the group-specific estimates were also imprecise. Therefore, the fact that the group-specific point estimates were in the same direction does not allow us to conclude that the differences between groups are small, or to narrow down explanations such as isolation or childcare burden. Third, participants were recruited from an internet survey panel, and the analysis was restricted to survey waves in which participants were working as employees and had worked in the past month. Moreover, only 19,056 of the 39,773 participants were observed in two or more waves, and 4,879 of these participants changed their WFH frequency category during follow-up. Although participants observed in only one wave contributed to the between-person estimates, they did not contribute directly to identifying the within-person associations. Thus, the within-person estimates were primarily informed by a smaller subset of participants whose WFH frequency changed over time. Selection related to survey participation and employment status may also limit the generalizability of the findings.

## Conclusion

Among employees in Japan, WFH at low frequencies (2–3 times a month or once a week) was associated with higher psychological distress than no WFH, both between and within persons. This was also the case for WFH 2–3 times a week. These results suggest the need to pay attention to the mental health of employees who work from home only a few times a month or a week.

## Statements and Declarations

### Ethics approval

The JACSIS study was approved by the Research Ethics Committee of the Osaka International Cancer Institute on 19 June 2020, before the first survey (approval number 20084). This study was approved by the Research Ethics Committees of Tohoku University Graduate School of Medicine (approval number 2025-1-523) and Hokkaido University Hospital (approval number 025-0281).

### Consent to participate

Informed consent was obtained electronically from all participants at the time of data collection.

### Consent to publish

Not applicable. This study is a secondary analysis of survey data, and the manuscript contains no information that could identify individual participants.

### Data availability

The data used in this study are not deposited in a public repository because they contain personally identifiable or potentially sensitive information. In accordance with the ethical guidelines in Japan, restrictions on data dissemination were imposed by the Research Ethics Committee of the Osaka International Cancer Institute and the Research Ethics Committee of Tohoku University Graduate School of Medicine.

Information on data access is available on the JACSIS website (https://jacsis-study.jp), and all data inquiries will be channeled through the JACSIS research office.

### Code availability

The R code used for the analyses is available on reasonable request. **Author contributions** SF contributed to the study conception and design. SF was responsible for data integrity, performed the statistical analysis, and drafted the manuscript. HS contributed to the interpretation of the results and revised the manuscript. SI supervised the study, contributed to the interpretation of the results, and provided administrative and technical support. TT conducted the JACSIS survey and managed the data. All authors critically reviewed the manuscript, approved the final version, and accept responsibility for the decision to submit it for publication.

## Supplementary information

The Supplementary Material contains the following.

Supplementary Methods: definitions of covariates; analysis of K6 at the next survey wave

Table S1: Characteristics of the analytic sample by survey wave

Table S2: Characteristics of participants at their first wave in the analysis, by frequency of working from home (all characteristics)

Table S3: All coefficients of the main within–between models (K6 score, K6 ≥5, and K6 ≥13)

Table S4: Proportion of person-waves in 2021–2024 with K6 observed at the next survey wave, by characteristics at the current wave

Table S5: Characteristics at the current wave of the pairs used in the analysis of K6 at the next survey wave, by frequency of working from home at the current wave

Table S6: Frequency of working from home by sex and by household circumstances at the first wave in the analysis

Fig. S1: Selection of participants

Fig. S2: Frequency of working from home over the survey waves (numbers of participants in each wave and transitions between adjacent waves)

Fig. S3: Frequency of working from home at the current wave and K6 at the next survey wave (models with and without adjustment for K6 at the current wave)

Fig. S4: Supplementary analyses (adjustment for basic characteristics only, restriction to ≥30 hours a week, restriction to mainly desk work, and weighting by the inverse of the number of waves)

