## Supplementary Material for "Working from home a few times a month to a few times a week is associated with higher psychological distress: between- and within-person analyses of a five-wave panel of employees in Japan"

#### Supplementary Methods

##### 1. Definitions of covariates

Sex, age, region of residence, and education were fixed at the values from each person's first response in the 2021–2025 surveys. The other covariates were measured at each wave.

| Variable | Time point | Categories |
|---|---|---|
| Sex | First response in 2021–2025 | Men; women |
| Age | First response (converted to the 2021 equivalent) | <20, 20–29, 30–39, 40–49, 50–59, 60–69, and ≥70 years |
| Region of residence | First response | Hokkaido; Tohoku; Kanto; Chubu; Kinki; Chugoku; Shikoku; Kyushu/Okinawa |
| Education | First response | High school or less; vocational school, junior college, or college of technology; university or higher; other |
| Occupation | Each wave | Professional or technical; clerical; sales; service; security; production (including machine assembly); transport or machine operation; construction or mining; carrying, cleaning or packaging; other |
| Industry | Each wave | Agriculture, forestry and fisheries; mining; construction; manufacturing; electricity, gas, heat supply and water; information and communications; transport; wholesale trade; retail trade; finance and insurance; real estate; eating and drinking places (serving alcohol); eating and drinking places (not serving alcohol); accommodations; medical care (at hospitals or clinics); medical care (not at hospitals or clinics); welfare; education and learning support; other services or public service |
| Employment type | Each wave | Regular staff or employee (including managers); dispatched worker; contract or entrusted employee; part-time or temporary worker (including students doing part-time work) |
| Company size | Each wave | 1–4, 5–99, 100–999, and ≥1,000 employees; don't know |
| Household type | Each wave | Living alone; living with a spouse or partner and no child <18 years; living with a spouse or partner and a child <18 years; other |
| Annual household income | Each wave | <2, 2 to <4, 4 to <6, 6 to <8, 8 to <10, and ≥10 million JPY; prefer not to answer; don't know |
| Survey wave | Each wave | 2021; 2022; 2023; 2024; 2025 |

##### 2. Analysis of K6 at the next survey wave

We defined a pair as the same person's responses in two adjacent surveys (the current wave and the next wave). The current waves were the person-waves in the analytic sample in 2021–2024; one person could contribute up to four pairs. If a person did not take part in an intermediate survey, the responses before and after that survey were not paired. Pairs with K6 observed at the next survey wave were used in the analysis, regardless of employment status at the next wave.

The outcome was the K6 score at the next survey wave, and the exposure was the frequency of working from home (WFH) at the current wave (seven categories). We adjusted for K6 at the current wave (natural spline with internal knots at 5 and 13), the same covariates as in the main analysis (values at the current wave), sex, age, region, education, and the pair of survey waves. Because the same person could contribute several pairs, standard errors were clustered by person. A model without adjustment for K6 at the current wave was also estimated using the same pairs. Because the interval between surveys was not constant, the estimates do not represent changes per year.

The estimates represent differences in distress at the next survey wave among people with similar levels of distress at the current wave. K6 at the current wave may itself be associated with earlier WFH. In addition, the frequency of WFH may change before the next survey, and the people who respond to the next survey may be a selected group. The proportion of person-waves in 2021–2024 with K6 observed at the next survey wave is shown in Table S4, and the characteristics at the current wave of the pairs used in the analysis are shown in Table S5.

### Supplementary Tables

#### Table S1. Characteristics of the analytic sample by survey wave

| Characteristic | All waves | 2021 | 2022 | 2023 | 2024 | 2025 |
|---|---|---|---|---|---|---|
| Survey period | 2021–2026 | Sep–Oct 2021 | Sep–Oct 2022 | Sep–Nov 2023 | Dec 2024–Jan 2025 | Dec 2025–Feb 2026 |
| Participants, n | 78,498 | 15,167 | 16,259 | 16,156 | 14,354 | 16,562 |
| **Sex** | | | | | | |
| Men | 41,765 (53.2) | 8,290 (54.7) | 8,816 (54.2) | 8,575 (53.1) | 7,718 (53.8) | 8,366 (50.5) |
| Women | 36,733 (46.8) | 6,877 (45.3) | 7,443 (45.8) | 7,581 (46.9) | 6,636 (46.2) | 8,196 (49.5) |
| **Age group (2021 equivalent), years** | | | | | | |
| <20 | 3,099 (3.9) | 306 (2.0) | 341 (2.1) | 848 (5.2) | 618 (4.3) | 986 (6.0) |
| 20–29 | 19,004 (24.2) | 2,727 (18.0) | 3,667 (22.6) | 4,468 (27.7) | 3,529 (24.6) | 4,613 (27.9) |
| 30–39 | 17,351 (22.1) | 3,036 (20.0) | 4,494 (27.6) | 3,144 (19.5) | 3,030 (21.1) | 3,647 (22.0) |
| 40–49 | 16,891 (21.5) | 3,887 (25.6) | 3,389 (20.8) | 3,239 (20.0) | 3,129 (21.8) | 3,247 (19.6) |
| 50–59 | 13,384 (17.1) | 3,030 (20.0) | 2,541 (15.6) | 2,661 (16.5) | 2,573 (17.9) | 2,579 (15.6) |
| 60–69 | 7,087 (9.0) | 1,717 (11.3) | 1,444 (8.9) | 1,462 (9.0) | 1,226 (8.5) | 1,238 (7.5) |
| ≥70 | 1,682 (2.1) | 464 (3.1) | 383 (2.4) | 334 (2.1) | 249 (1.7) | 252 (1.5) |
| **Education** | | | | | | |
| High school or less | 17,634 (22.5) | 3,581 (23.6) | 3,574 (22.0) | 3,664 (22.7) | 3,249 (22.6) | 3,566 (21.5) |
| Vocational/junior college | 15,947 (20.3) | 3,249 (21.4) | 3,322 (20.4) | 3,229 (20.0) | 2,907 (20.3) | 3,240 (19.6) |
| University or higher | 44,487 (56.7) | 8,254 (54.4) | 9,263 (57.0) | 9,181 (56.8) | 8,118 (56.6) | 9,671 (58.4) |
| Other | 430 (0.5) | 83 (0.5) | 100 (0.6) | 82 (0.5) | 80 (0.6) | 85 (0.5) |
| **Region** | | | | | | |
| Hokkaido | 3,331 (4.2) | 606 (4.0) | 659 (4.1) | 698 (4.3) | 620 (4.3) | 748 (4.5) |
| Tohoku | 4,723 (6.0) | 979 (6.5) | 960 (5.9) | 953 (5.9) | 880 (6.1) | 951 (5.7) |
| Kanto | 29,624 (37.7) | 5,463 (36.0) | 5,994 (36.9) | 6,320 (39.1) | 5,500 (38.3) | 6,347 (38.3) |
| Chubu | 13,173 (16.8) | 2,591 (17.1) | 2,723 (16.7) | 2,676 (16.6) | 2,406 (16.8) | 2,777 (16.8) |
| Kinki | 14,339 (18.3) | 2,650 (17.5) | 3,004 (18.5) | 2,918 (18.1) | 2,635 (18.4) | 3,132 (18.9) |
| Chugoku | 4,272 (5.4) | 853 (5.6) | 921 (5.7) | 847 (5.2) | 768 (5.4) | 883 (5.3) |
| Shikoku | 2,003 (2.6) | 424 (2.8) | 438 (2.7) | 382 (2.4) | 351 (2.4) | 408 (2.5) |
| Kyushu/Okinawa | 7,033 (9.0) | 1,601 (10.6) | 1,560 (9.6) | 1,362 (8.4) | 1,194 (8.3) | 1,316 (7.9) |
| **Employment type** | | | | | | |
| Regular employee | 51,081 (65.1) | 9,625 (63.5) | 10,859 (66.8) | 10,396 (64.3) | 9,406 (65.5) | 10,795 (65.2) |
| Dispatched worker | 2,109 (2.7) | 396 (2.6) | 392 (2.4) | 445 (2.8) | 403 (2.8) | 473 (2.9) |
| Contract or entrusted employee | 5,613 (7.2) | 1,092 (7.2) | 1,152 (7.1) | 1,155 (7.1) | 1,072 (7.5) | 1,142 (6.9) |
| Part-time or temporary worker | 19,695 (25.1) | 4,054 (26.7) | 3,856 (23.7) | 4,160 (25.7) | 3,473 (24.2) | 4,152 (25.1) |
| **Occupation** | | | | | | |
| Professional or technical | 18,364 (23.4) | 3,470 (22.9) | 3,980 (24.5) | 3,725 (23.1) | 3,349 (23.3) | 3,840 (23.2) |
| Clerical | 21,337 (27.2) | 4,035 (26.6) | 4,350 (26.8) | 4,318 (26.7) | 3,965 (27.6) | 4,669 (28.2) |
| Sales | 9,846 (12.5) | 1,964 (12.9) | 2,063 (12.7) | 2,021 (12.5) | 1,756 (12.2) | 2,042 (12.3) |
| Service | 6,419 (8.2) | 1,183 (7.8) | 1,254 (7.7) | 1,377 (8.5) | 1,178 (8.2) | 1,427 (8.6) |
| Security | 1,072 (1.4) | 191 (1.3) | 227 (1.4) | 238 (1.5) | 205 (1.4) | 211 (1.3) |
| Production | 4,623 (5.9) | 950 (6.3) | 999 (6.1) | 893 (5.5) | 830 (5.8) | 951 (5.7) |
| Transport or machine operation | 1,368 (1.7) | 265 (1.7) | 291 (1.8) | 299 (1.9) | 259 (1.8) | 254 (1.5) |
| Construction or mining | 824 (1.0) | 150 (1.0) | 187 (1.2) | 170 (1.1) | 146 (1.0) | 171 (1.0) |
| Carrying, cleaning or packaging | 2,475 (3.2) | 461 (3.0) | 466 (2.9) | 501 (3.1) | 476 (3.3) | 571 (3.4) |
| Other | 12,170 (15.5) | 2,498 (16.5) | 2,442 (15.0) | 2,614 (16.2) | 2,190 (15.3) | 2,426 (14.6) |
| **Company size, employees** | | | | | | |
| 1–4 | 2,636 (3.4) | 569 (3.8) | 541 (3.3) | 528 (3.3) | 470 (3.3) | 528 (3.2) |

| Characteristic | All waves | 2021 | 2022 | 2023 | 2024 | 2025 |
|---|---|---|---|---|---|---|
| 5–99 | 23,819 (30.3) | 4,786 (31.6) | 4,861 (29.9) | 4,877 (30.2) | 4,316 (30.1) | 4,979 (30.1) |
| 100–999 | 20,800 (26.5) | 3,990 (26.3) | 4,365 (26.8) | 4,276 (26.5) | 3,742 (26.1) | 4,427 (26.7) |
| ≥1,000 | 23,290 (29.7) | 4,289 (28.3) | 4,935 (30.4) | 4,754 (29.4) | 4,272 (29.8) | 5,040 (30.4) |
| Don't know | 7,953 (10.1) | 1,533 (10.1) | 1,557 (9.6) | 1,721 (10.7) | 1,554 (10.8) | 1,588 (9.6) |
| Working ≥30 hours a week | 60,171 (76.7) | 11,559 (76.2) | 12,497 (76.9) | 12,175 (75.4) | 11,134 (77.6) | 12,806 (77.3) |
| Mainly desk work | 36,896 (47.0) | 7,013 (46.2) | 7,579 (46.6) | 7,509 (46.5) | 6,863 (47.8) | 7,932 (47.9) |
| **Household type** | | | | | | |
| Living alone | 19,592 (25.0) | 3,572 (23.6) | 3,650 (22.4) | 4,267 (26.4) | 3,630 (25.3) | 4,473 (27.0) |
| With spouse or partner, no child <18 | 21,836 (27.8) | 4,497 (29.6) | 4,145 (25.5) | 4,562 (28.2) | 4,103 (28.6) | 4,529 (27.3) |
| With spouse or partner and child <18 | 19,139 (24.4) | 3,539 (23.3) | 4,922 (30.3) | 3,385 (21.0) | 3,452 (24.0) | 3,841 (23.2) |
| Other | 17,931 (22.8) | 3,559 (23.5) | 3,542 (21.8) | 3,942 (24.4) | 3,169 (22.1) | 3,719 (22.5) |
| **Annual household income, million JPY** | | | | | | |
| <2 | 4,222 (5.4) | 838 (5.5) | 833 (5.1) | 867 (5.4) | 758 (5.3) | 926 (5.6) |
| 2 to <4 | 12,855 (16.4) | 2,606 (17.2) | 2,644 (16.3) | 2,803 (17.3) | 2,211 (15.4) | 2,591 (15.6) |
| 4 to <6 | 15,823 (20.2) | 3,160 (20.8) | 3,390 (20.8) | 3,239 (20.0) | 2,857 (19.9) | 3,177 (19.2) |
| 6 to <8 | 12,307 (15.7) | 2,422 (16.0) | 2,706 (16.6) | 2,470 (15.3) | 2,139 (14.9) | 2,570 (15.5) |
| 8 to <10 | 8,834 (11.3) | 1,674 (11.0) | 1,929 (11.9) | 1,734 (10.7) | 1,591 (11.1) | 1,906 (11.5) |
| ≥10 | 9,728 (12.4) | 1,703 (11.2) | 1,984 (12.2) | 1,829 (11.3) | 1,831 (12.8) | 2,381 (14.4) |
| Prefer not to answer | 6,967 (8.9) | 1,396 (9.2) | 1,305 (8.0) | 1,460 (9.0) | 1,438 (10.0) | 1,368 (8.3) |
| Don't know | 7,762 (9.9) | 1,368 (9.0) | 1,468 (9.0) | 1,754 (10.9) | 1,529 (10.7) | 1,643 (9.9) |
| K6 score, mean (SD) | 4.9 (5.7) | 4.6 (5.5) | 4.8 (5.7) | 4.9 (5.8) | 5.3 (5.9) | 5.0 (5.8) |
| **Frequency of working from home** | | | | | | |
| None | 61,706 (78.6) | 11,500 (75.8) | 12,570 (77.3) | 13,015 (80.6) | 11,403 (79.4) | 13,218 (79.8) |
| Once a month | 2,367 (3.0) | 419 (2.8) | 505 (3.1) | 461 (2.9) | 447 (3.1) | 535 (3.2) |
| 2–3 times a month | 2,595 (3.3) | 576 (3.8) | 547 (3.4) | 484 (3.0) | 475 (3.3) | 513 (3.1) |
| Once a week | 2,916 (3.7) | 582 (3.8) | 633 (3.9) | 530 (3.3) | 568 (4.0) | 603 (3.6) |
| 2–3 times a week | 4,279 (5.5) | 1,001 (6.6) | 928 (5.7) | 813 (5.0) | 715 (5.0) | 822 (5.0) |
| 4–5 times a week | 2,565 (3.3) | 594 (3.9) | 626 (3.9) | 460 (2.8) | 405 (2.8) | 480 (2.9) |
| Almost daily | 2,070 (2.6) | 495 (3.3) | 450 (2.8) | 393 (2.4) | 341 (2.4) | 391 (2.4) |
| **K6 score category** | | | | | | |
| 0–4 | 46,798 (59.6) | 9,272 (61.1) | 9,834 (60.5) | 9,748 (60.3) | 8,144 (56.7) | 9,800 (59.2) |
| 5–12 | 22,449 (28.6) | 4,413 (29.1) | 4,537 (27.9) | 4,450 (27.5) | 4,264 (29.7) | 4,785 (28.9) |
| ≥13 | 9,251 (11.8) | 1,482 (9.8) | 1,888 (11.6) | 1,958 (12.1) | 1,946 (13.6) | 1,977 (11.9) |

Values are n (column %) of participants in each wave unless otherwise indicated. A person who took part in several waves is counted in each of those waves (78,498 person-waves from 39,773 persons).
Sex, age group, education and region were fixed at the first valid response in 2021–2025; age was converted to the 2021 equivalent.

**Table S2. Characteristics of participants at their first wave in the analysis, by frequency of working from home (all characteristics)**

| Characteristic | Total | None | Once a month | 2–3 times a month | Once a week | 2–3 times a week | 4–5 times a week | Almost daily |
|---|---|---|---|---|---|---|---|---|
| Persons, n | 39,773 | 30,877 | 1,187 | 1,396 | 1,543 | 2,256 | 1,353 | 1,161 |
| **Sex** | | | | | | | | |
| Men | 20,058 (50.4) | 14,538 (47.1) | 785 (66.1) | 923 (66.1) | 1,016 (65.8) | 1,358 (60.2) | 794 (58.7) | 644 (55.5) |
| Women | 19,715 (49.6) | 16,339 (52.9) | 402 (33.9) | 473 (33.9) | 527 (34.2) | 898 (39.8) | 559 (41.3) | 517 (44.5) |
| **Age group (2021 equivalent), years** | | | | | | | | |
| <20 | 2,357 (5.9) | 2,013 (6.5) | 48 (4.0) | 65 (4.7) | 79 (5.1) | 87 (3.9) | 36 (2.7) | 29 (2.5) |
| 20–29 | 11,573 (29.1) | 8,652 (28.0) | 414 (34.9) | 488 (35.0) | 577 (37.4) | 707 (31.3) | 389 (28.8) | 346 (29.8) |
| 30–39 | 9,032 (22.7) | 6,816 (22.1) | 325 (27.4) | 334 (23.9) | 373 (24.2) | 543 (24.1) | 356 (26.3) | 285 (24.5) |
| 40–49 | 7,361 (18.5) | 5,719 (18.5) | 189 (15.9) | 263 (18.8) | 242 (15.7) | 422 (18.7) | 264 (19.5) | 262 (22.6) |
| 50–59 | 5,686 (14.3) | 4,504 (14.6) | 153 (12.9) | 169 (12.1) | 178 (11.5) | 310 (13.7) | 219 (16.2) | 153 (13.2) |
| 60–69 | 3,018 (7.6) | 2,527 (8.2) | 47 (4.0) | 62 (4.4) | 80 (5.2) | 164 (7.3) | 77 (5.7) | 61 (5.3) |
| ≥70 | 746 (1.9) | 646 (2.1) | 11 (0.9) | 15 (1.1) | 14 (0.9) | 23 (1.0) | 12 (0.9) | 25 (2.2) |
| **Education** | | | | | | | | |
| High school or less | 8,900 (22.4) | 7,911 (25.6) | 135 (11.4) | 176 (12.6) | 182 (11.8) | 217 (9.6) | 134 (9.9) | 145 (12.5) |
| Vocational/junior college | 8,075 (20.3) | 6,945 (22.5) | 143 (12.0) | 167 (12.0) | 202 (13.1) | 250 (11.1) | 177 (13.1) | 191 (16.5) |
| University or higher | 22,561 (56.7) | 15,844 (51.3) | 906 (76.3) | 1,048 (75.1) | 1,151 (74.6) | 1,772 (78.5) | 1,033 (76.3) | 807 (69.5) |
| Other | 237 (0.6) | 177 (0.6) | 3 (0.3) | 5 (0.4) | 8 (0.5) | 17 (0.8) | 9 (0.7) | 18 (1.6) |
| **Region** | | | | | | | | |
| Hokkaido | 1,671 (4.2) | 1,434 (4.6) | 40 (3.4) | 49 (3.5) | 44 (2.9) | 49 (2.2) | 19 (1.4) | 36 (3.1) |
| Tohoku | 2,369 (6.0) | 2,061 (6.7) | 47 (4.0) | 56 (4.0) | 53 (3.4) | 77 (3.4) | 31 (2.3) | 44 (3.8) |
| Kanto | 15,181 (38.2) | 10,315 (33.4) | 507 (42.7) | 647 (46.3) | 778 (50.4) | 1,318 (58.4) | 915 (67.6) | 701 (60.4) |
| Chubu | 6,617 (16.6) | 5,552 (18.0) | 198 (16.7) | 190 (13.6) | 210 (13.6) | 240 (10.6) | 112 (8.3) | 115 (9.9) |
| Kinki | 7,307 (18.4) | 5,773 (18.7) | 229 (19.3) | 267 (19.1) | 284 (18.4) | 390 (17.3) | 194 (14.3) | 170 (14.6) |
| Chugoku | 2,160 (5.4) | 1,889 (6.1) | 50 (4.2) | 62 (4.4) | 53 (3.4) | 59 (2.6) | 19 (1.4) | 28 (2.4) |
| Shikoku | 996 (2.5) | 900 (2.9) | 26 (2.2) | 21 (1.5) | 25 (1.6) | 11 (0.5) | 5 (0.4) | 8 (0.7) |
| Kyushu/Okinawa | 3,472 (8.7) | 2,953 (9.6) | 90 (7.6) | 104 (7.4) | 96 (6.2) | 112 (5.0) | 58 (4.3) | 59 (5.1) |
| **Employment type** | | | | | | | | |
| Regular employee | 25,185 (63.3) | 18,100 (58.6) | 983 (82.8) | 1,114 (79.8) | 1,211 (78.5) | 1,825 (80.9) | 1,051 (77.7) | 901 (77.6) |
| Dispatched worker | 1,055 (2.7) | 846 (2.7) | 14 (1.2) | 25 (1.8) | 27 (1.7) | 51 (2.3) | 55 (4.1) | 37 (3.2) |
| Contract or entrusted employee | 2,455 (6.2) | 1,947 (6.3) | 46 (3.9) | 63 (4.5) | 88 (5.7) | 133 (5.9) | 96 (7.1) | 82 (7.1) |
| Part-time or temporary worker | 11,078 (27.9) | 9,984 (32.3) | 144 (12.1) | 194 (13.9) | 217 (14.1) | 247 (10.9) | 151 (11.2) | 141 (12.1) |
| **Occupation** | | | | | | | | |
| Professional or technical | 9,102 (22.9) | 6,373 (20.6) | 360 (30.3) | 379 (27.1) | 364 (23.6) | 658 (29.2) | 519 (38.4) | 449 (38.7) |
| Clerical | 10,259 (25.8) | 7,349 (23.8) | 384 (32.4) | 446 (31.9) | 498 (32.3) | 807 (35.8) | 446 (33.0) | 329 (28.3) |
| Sales | 5,037 (12.7) | 3,682 (11.9) | 192 (16.2) | 233 (16.7) | 284 (18.4) | 354 (15.7) | 151 (11.2) | 141 (12.1) |
| Service | 3,545 (8.9) | 3,205 (10.4) | 38 (3.2) | 66 (4.7) | 100 (6.5) | 64 (2.8) | 34 (2.5) | 38 (3.3) |
| Security | 504 (1.3) | 455 (1.5) | 2 (0.2) | 7 (0.5) | 15 (1.0) | 16 (0.7) | 8 (0.6) | 1 (0.1) |
| Production | 2,231 (5.6) | 2,050 (6.6) | 34 (2.9) | 49 (3.5) | 41 (2.7) | 33 (1.5) | 12 (0.9) | 12 (1.0) |
| Transport or machine operation | 671 (1.7) | 606 (2.0) | 14 (1.2) | 12 (0.9) | 16 (1.0) | 15 (0.7) | 2 (0.1) | 6 (0.5) |
| Construction or mining | 430 (1.1) | 351 (1.1) | 12 (1.0) | 13 (0.9) | 24 (1.6) | 15 (0.7) | 7 (0.5) | 8 (0.7) |
| Carrying, cleaning or packaging | 1,227 (3.1) | 1,142 (3.7) | 8 (0.7) | 19 (1.4) | 19 (1.2) | 20 (0.9) | 11 (0.8) | 8 (0.7) |
| Other | 6,767 (17.0) | 5,664 (18.3) | 143 (12.0) | 172 (12.3) | 182 (11.8) | 274 (12.1) | 163 (12.0) | 169 (14.6) |
| **Company size, employees** | | | | | | | | |
| 1–4 | 1,341 (3.4) | 1,089 (3.5) | 25 (2.1) | 35 (2.5) | 33 (2.1) | 57 (2.5) | 33 (2.4) | 69 (5.9) |
| 5–99 | 12,339 (31.0) | 10,537 (34.1) | 227 (19.1) | 331 (23.7) | 329 (21.3) | 416 (18.4) | 236 (17.4) | 263 (22.7) |
| 100–999 | 10,279 (25.8) | 7,926 (25.7) | 303 (25.5) | 366 (26.2) | 462 (29.9) | 603 (26.7) | 350 (25.9) | 269 (23.2) |
| ≥1,000 | 11,486 (28.9) | 7,539 (24.4) | 573 (48.3) | 600 (43.0) | 621 (40.2) | 1,041 (46.1) | 653 (48.3) | 459 (39.5) |

| Characteristic | Total | None | Once a month | 2–3 times a month | Once a week | 2–3 times a week | 4–5 times a week | Almost daily |
|---|---|---|---|---|---|---|---|---|
| Don't know | 4,328 (10.9) | 3,786 (12.3) | 59 (5.0) | 64 (4.6) | 98 (6.4) | 139 (6.2) | 81 (6.0) | 101 (8.7) |
| Working ≥30 hours a week | 29,492 (74.2) | 22,030 (71.3) | 1,019 (85.8) | 1,148 (82.2) | 1,229 (79.7) | 1,915 (84.9) | 1,179 (87.1) | 972 (83.7) |
| Mainly desk work | 17,652 (44.4) | 11,179 (36.2) | 754 (63.5) | 920 (65.9) | 997 (64.6) | 1,722 (76.3) | 1,130 (83.5) | 950 (81.8) |
| **Household type** | | | | | | | | |
| Living alone | 10,138 (25.5) | 7,645 (24.8) | 309 (26.0) | 383 (27.4) | 437 (28.3) | 664 (29.4) | 364 (26.9) | 336 (28.9) |
| With spouse or partner, no child <18 | 9,869 (24.8) | 7,678 (24.9) | 286 (24.1) | 322 (23.1) | 364 (23.6) | 591 (26.2) | 340 (25.1) | 288 (24.8) |
| With spouse or partner and child <18 | 9,809 (24.7) | 7,312 (23.7) | 361 (30.4) | 428 (30.7) | 417 (27.0) | 611 (27.1) | 385 (28.5) | 295 (25.4) |
| Other | 9,957 (25.0) | 8,242 (26.7) | 231 (19.5) | 263 (18.8) | 325 (21.1) | 390 (17.3) | 264 (19.5) | 242 (20.8) |
| **Annual household income, million JPY** | | | | | | | | |
| <2 | 2,375 (6.0) | 2,062 (6.7) | 29 (2.4) | 56 (4.0) | 69 (4.5) | 74 (3.3) | 36 (2.7) | 49 (4.2) |
| 2 to <4 | 6,717 (16.9) | 5,641 (18.3) | 137 (11.5) | 175 (12.5) | 204 (13.2) | 258 (11.4) | 163 (12.0) | 139 (12.0) |
| 4 to <6 | 7,995 (20.1) | 6,275 (20.3) | 231 (19.5) | 302 (21.6) | 302 (19.6) | 426 (18.9) | 247 (18.3) | 212 (18.3) |
| 6 to <8 | 6,115 (15.4) | 4,653 (15.1) | 227 (19.1) | 237 (17.0) | 246 (15.9) | 381 (16.9) | 185 (13.7) | 186 (16.0) |
| 8 to <10 | 4,304 (10.8) | 3,073 (10.0) | 179 (15.1) | 191 (13.7) | 228 (14.8) | 287 (12.7) | 199 (14.7) | 147 (12.7) |
| ≥10 | 4,687 (11.8) | 2,862 (9.3) | 223 (18.8) | 266 (19.1) | 296 (19.2) | 525 (23.3) | 307 (22.7) | 208 (17.9) |
| Prefer not to answer | 3,394 (8.5) | 2,741 (8.9) | 65 (5.5) | 80 (5.7) | 100 (6.5) | 173 (7.7) | 124 (9.2) | 111 (9.6) |
| Don't know | 4,186 (10.5) | 3,570 (11.6) | 96 (8.1) | 89 (6.4) | 98 (6.4) | 132 (5.9) | 92 (6.8) | 109 (9.4) |
| K6 score, mean (SD) | 5.1 (5.7) | 4.9 (5.7) | 5.3 (5.7) | 6.3 (5.9) | 6.4 (5.8) | 5.2 (5.7) | 5.0 (5.6) | 5.4 (6.1) |
| **K6 score category** | | | | | | | | |
| 0–4 | 23,055 (58.0) | 18,302 (59.3) | 673 (56.7) | 677 (48.5) | 700 (45.4) | 1,266 (56.1) | 790 (58.4) | 647 (55.7) |
| 5–12 | 11,908 (29.9) | 9,023 (29.2) | 355 (29.9) | 476 (34.1) | 575 (37.3) | 717 (31.8) | 400 (29.6) | 362 (31.2) |
| ≥13 | 4,810 (12.1) | 3,552 (11.5) | 159 (13.4) | 243 (17.4) | 268 (17.4) | 273 (12.1) | 163 (12.0) | 152 (13.1) |
| **Number of waves in the analysis** | | | | | | | | |
| 1 | 20,717 (52.1) | 16,230 (52.6) | 610 (51.4) | 722 (51.7) | 811 (52.6) | 1,097 (48.6) | 664 (49.1) | 583 (50.2) |
| 2 | 7,714 (19.4) | 6,027 (19.5) | 227 (19.1) | 268 (19.2) | 280 (18.1) | 439 (19.5) | 228 (16.9) | 245 (21.1) |
| 3 | 5,396 (13.6) | 4,104 (13.3) | 164 (13.8) | 192 (13.8) | 235 (15.2) | 329 (14.6) | 206 (15.2) | 166 (14.3) |
| 4 | 3,565 (9.0) | 2,731 (8.8) | 121 (10.2) | 128 (9.2) | 124 (8.0) | 220 (9.8) | 141 (10.4) | 100 (8.6) |
| 5 | 2,381 (6.0) | 1,785 (5.8) | 65 (5.5) | 86 (6.2) | 93 (6.0) | 171 (7.6) | 114 (8.4) | 67 (5.8) |

Values are n (column %) unless otherwise indicated. Columns show the frequency of working from home at the first wave in the analysis. Table 1 in the main text shows a subset of these characteristics.
Sex, age group, education and region were fixed at the first valid response in 2021–2025; age was converted to the 2021 equivalent. The other characteristics were taken from the first wave in the analysis.

**Table S3. All coefficients of the main within–between models**

| Term | K6 score, points | K6 ≥5, percentage points | K6 ≥13, percentage points |
|---|---|---|---|
| **Within-person terms (deviation from the person mean)** | | | |
| **Frequency of working from home (reference: None)** | | | |
| Once a month | 0.07 (−0.17, 0.30) | 1.1 (−1.2, 3.4) | 0.5 (−1.2, 2.3) |
| 2–3 times a month | 0.53 (0.28, 0.77) | 3.5 (1.2, 5.7) | 2.7 (0.7, 4.6) |
| Once a week | 0.45 (0.20, 0.69) | 4.6 (2.3, 6.9) | 1.5 (−0.4, 3.4) |
| 2–3 times a week | 0.35 (0.10, 0.60) | 3.8 (1.6, 6.0) | 2.1 (0.3, 4.0) |
| 4–5 times a week | 0.03 (−0.28, 0.34) | 0.9 (−2.0, 3.8) | 0.4 (−2.0, 2.9) |
| Almost daily | 0.20 (−0.16, 0.56) | 1.9 (−1.3, 5.1) | 1.0 (−1.6, 3.6) |
| **Occupation (reference: Professional or technical)** | | | |
| Clerical | 0.01 (−0.22, 0.23) | −0.3 (−2.5, 1.9) | 0.3 (−1.3, 1.9) |
| Sales | 0.22 (−0.04, 0.49) | 3.2 (0.7, 5.8) | 0.7 (−1.2, 2.7) |
| Service | −0.02 (−0.27, 0.23) | −0.8 (−3.2, 1.6) | 0.3 (−1.6, 2.1) |
| Security | 0.52 (−0.08, 1.11) | 3.8 (−1.6, 9.2) | 2.6 (−1.4, 6.6) |
| Production | −0.02 (−0.39, 0.34) | 0.0 (−3.3, 3.3) | −1.3 (−4.0, 1.5) |
| Transport or machine operation | 0.09 (−0.40, 0.58) | −1.7 (−6.2, 2.8) | 3.0 (−0.5, 6.5) |
| Construction or mining | 0.50 (−0.09, 1.10) | 3.7 (−1.7, 9.2) | 2.3 (−1.8, 6.3) |
| Carrying, cleaning or packaging | 0.11 (−0.25, 0.48) | −0.7 (−4.0, 2.7) | 1.7 (−0.9, 4.3) |
| Other | 0.15 (−0.04, 0.34) | 0.7 (−1.2, 2.5) | 1.1 (−0.3, 2.5) |
| **Industry (reference: Agriculture, forestry and fisheries)** | | | |
| Mining | 1.57 (−0.14, 3.27) | 10.2 (−6.4, 26.8) | 2.4 (−16.8, 21.5) |
| Construction | 1.13 (−0.08, 2.34) | 6.3 (−6.2, 18.7) | 9.9 (0.7, 19.2) |
| Manufacturing | 0.88 (−0.25, 2.01) | 5.3 (−6.6, 17.2) | 6.4 (−2.1, 14.9) |
| Electricity, gas, heat supply and water | 0.81 (−0.47, 2.09) | 2.7 (−10.3, 15.7) | 9.0 (−0.9, 18.8) |
| Information and communications | 0.62 (−0.54, 1.79) | 3.8 (−8.3, 15.9) | 4.1 (−4.7, 13.0) |
| Transport | 0.65 (−0.53, 1.82) | 4.5 (−7.8, 16.7) | 3.3 (−5.6, 12.2) |
| Wholesale trade | 0.61 (−0.56, 1.79) | 4.8 (−7.5, 17.1) | 4.7 (−4.1, 13.6) |
| Retail trade | 0.63 (−0.51, 1.78) | 4.6 (−7.3, 16.5) | 4.2 (−4.6, 12.9) |
| Finance and insurance | 0.52 (−0.74, 1.77) | 6.0 (−6.9, 18.9) | 4.6 (−5.1, 14.3) |
| Real estate | 0.77 (−0.56, 2.10) | 7.3 (−6.2, 20.7) | 2.1 (−7.8, 12.0) |
| Eating and drinking places (serving alcohol) | 0.44 (−0.80, 1.68) | 3.8 (−9.0, 16.6) | 1.9 (−7.6, 11.4) |
| Eating and drinking places (not serving alcohol) | 0.52 (−0.69, 1.73) | 6.3 (−6.3, 18.9) | 2.4 (−6.7, 11.5) |
| Accommodations | 0.05 (−1.33, 1.43) | 0.1 (−13.4, 13.6) | 3.7 (−7.1, 14.4) |
| Medical care (at hospitals or clinics) | 0.37 (−0.80, 1.55) | 4.3 (−8.0, 16.6) | 4.2 (−4.8, 13.2) |
| Medical care (not at hospitals or clinics) | 0.45 (−0.73, 1.63) | 3.8 (−8.5, 16.1) | 5.1 (−3.9, 14.1) |
| Welfare | 0.24 (−0.93, 1.40) | 1.7 (−10.3, 13.8) | 3.4 (−5.6, 12.3) |
| Education and learning support | 0.41 (−0.74, 1.56) | 3.8 (−8.2, 15.8) | 3.9 (−4.8, 12.5) |
| Other services or public service | 0.61 (−0.50, 1.73) | 3.4 (−8.2, 15.1) | 4.2 (−4.2, 12.7) |
| **Employment type (reference: Regular employee)** | | | |
| Dispatched worker | −0.12 (−0.59, 0.35) | −1.0 (−5.1, 3.0) | −2.4 (−5.6, 0.8) |
| Contract or entrusted employee | −0.24 (−0.51, 0.02) | −4.1 (−6.7, −1.5) | 0.1 (−1.6, 1.9) |
| Part-time or temporary worker | −0.17 (−0.44, 0.10) | −2.5 (−5.0, 0.1) | −0.6 (−2.6, 1.4) |
| **Company size, employees (reference: 1–4)** | | | |
| 5–99 | −0.50 (−0.87, −0.14) | −3.9 (−7.3, −0.5) | −1.8 (−4.4, 0.9) |
| 100–999 | −0.42 (−0.81, −0.03) | −3.3 (−7.0, 0.4) | −0.5 (−3.3, 2.4) |
| ≥1,000 | −0.42 (−0.83, −0.02) | −3.1 (−6.9, 0.6) | −1.0 (−3.9, 2.0) |
| Don't know | −0.46 (−0.86, −0.05) | −4.1 (−7.9, −0.4) | −0.7 (−3.6, 2.2) |
| **Household type (reference: Living alone)** | | | |
| With spouse or partner, no child <18 | 0.04 (−0.24, 0.31) | 0.7 (−1.8, 3.3) | −0.1 (−2.0, 1.8) |
| With spouse or partner and child <18 | −0.09 (−0.41, 0.23) | 0.1 (−2.9, 3.2) | −0.9 (−3.2, 1.3) |
| Other | −0.09 (−0.35, 0.17) | −0.1 (−2.4, 2.2) | −0.7 (−2.6, 1.3) |
| **Annual household income, million JPY (reference: <2)** | | | |
| 2 to <4 | −0.27 (−0.58, 0.03) | −1.9 (−4.6, 0.8) | −1.8 (−4.1, 0.5) |
| 4 to <6 | −0.45 (−0.78, −0.12) | −2.8 (−5.7, 0.1) | −3.2 (−5.6, −0.7) |
| 6 to <8 | −0.57 (−0.92, −0.22) | −5.2 (−8.3, −2.1) | −2.9 (−5.4, −0.4) |
| 8 to <10 | −0.70 (−1.07, −0.34) | −6.1 (−9.3, −2.9) | −3.5 (−6.1, −0.9) |
| ≥10 | −0.59 (−0.97, −0.20) | −4.2 (−7.6, −0.8) | −3.2 (−6.0, −0.4) |
| Prefer not to answer | −0.69 (−1.05, −0.33) | −5.0 (−8.2, −1.9) | −3.6 (−6.2, −1.0) |
| Don't know | −0.56 (−0.91, −0.21) | −5.0 (−8.2, −1.9) | −3.8 (−6.4, −1.2) |
| **Survey wave (reference: 2021)** | | | |
| 2022 | −0.03 (−0.11, 0.05) | −0.9 (−1.7, 0.0) | 1.1 (0.5, 1.7) |
| 2023 | −0.09 (−0.18, 0.01) | −1.9 (−2.9, −1.0) | 1.3 (0.6, 1.9) |
| 2024 | 0.27 (0.17, 0.37) | 1.4 (0.4, 2.4) | 2.2 (1.5, 2.9) |
| 2025 | −0.35 (−0.47, −0.24) | −3.2 (−4.4, −2.1) | −0.4 (−1.2, 0.4) |
| **Between-person terms (person mean)** | | | |
| **Frequency of working from home (reference: None)** | | | |
| Once a month | 0.31 (−0.08, 0.70) | 2.9 (−0.4, 6.2) | 1.6 (−0.6, 3.7) |
| 2–3 times a month | 1.99 (1.61, 2.37) | 15.9 (12.8, 18.9) | 9.2 (7.0, 11.5) |
| Once a week | 1.80 (1.45, 2.14) | 16.6 (13.6, 19.5) | 6.4 (4.4, 8.4) |
| 2–3 times a week | 0.67 (0.37, 0.96) | 5.7 (3.3, 8.2) | 2.3 (0.7, 3.9) |
| 4–5 times a week | 0.43 (0.06, 0.79) | 4.0 (1.0, 7.1) | 1.9 (−0.1, 3.9) |
| Almost daily | 0.73 (0.32, 1.13) | 5.2 (1.9, 8.4) | 2.6 (0.6, 4.7) |
| **Occupation (reference: Professional or technical)** | | | |
| Clerical | 0.31 (0.12, 0.50) | 1.9 (0.3, 3.4) | 1.7 (0.7, 2.7) |

| Term | K6 score, points | K6 ≥5, percentage points | K6 ≥13, percentage points |
|---|---|---|---|
| Sales | −0.03 (−0.26, 0.20) | 0.5 (−1.5, 2.5) | −0.6 (−1.8, 0.6) |
| Service | 0.26 (−0.04, 0.56) | 2.8 (0.4, 5.2) | 0.6 (−1.1, 2.2) |
| Security | −0.40 (−0.95, 0.14) | −3.7 (−8.0, 0.7) | −0.9 (−3.6, 1.9) |
| Production | 0.75 (0.41, 1.09) | 6.6 (3.9, 9.4) | 3.1 (1.3, 4.9) |
| Transport or machine operation | 0.09 (−0.44, 0.62) | 0.5 (−3.9, 4.9) | 1.3 (−1.4, 4.0) |
| Construction or mining | −0.51 (−1.11, 0.09) | −3.4 (−8.6, 1.8) | −2.3 (−5.4, 0.8) |
| Carrying, cleaning or packaging | 0.52 (0.11, 0.94) | 4.2 (0.8, 7.6) | 1.7 (−0.5, 3.9) |
| Other | 0.01 (−0.21, 0.24) | 0.9 (−1.0, 2.7) | −0.3 (−1.5, 0.8) |
| **Industry (reference: Agriculture, forestry and fisheries)** | | | |
| Mining | −1.40 (−3.17, 0.38) | −9.2 (−25.7, 7.2) | −10.9 (−19.8, −2.0) |
| Construction | −0.56 (−1.48, 0.35) | −1.8 (−8.9, 5.3) | −5.9 (−11.1, −0.7) |
| Manufacturing | −0.82 (−1.70, 0.07) | −4.4 (−11.2, 2.5) | −7.6 (−12.7, −2.6) |
| Electricity, gas, heat supply and water | −0.41 (−1.42, 0.60) | −1.5 (−9.3, 6.3) | −4.9 (−10.7, 0.8) |
| Information and communications | −1.07 (−1.98, −0.15) | −6.3 (−13.4, 0.8) | −7.3 (−12.5, −2.0) |
| Transport | −0.88 (−1.81, 0.05) | −4.8 (−12.0, 2.4) | −7.3 (−12.5, −2.0) |
| Wholesale trade | −0.89 (−1.82, 0.04) | −4.9 (−12.2, 2.3) | −7.5 (−12.8, −2.3) |
| Retail trade | −0.50 (−1.39, 0.39) | −2.6 (−9.5, 4.3) | −5.2 (−10.3, −0.1) |
| Finance and insurance | −0.52 (−1.43, 0.40) | −1.4 (−8.5, 5.7) | −6.2 (−11.4, −0.9) |
| Real estate | −0.76 (−1.72, 0.19) | −3.5 (−11.0, 4.0) | −7.1 (−12.4, −1.7) |
| Eating and drinking places (serving alcohol) | −1.11 (−2.06, −0.16) | −6.3 (−13.7, 1.2) | −6.5 (−12.0, −1.1) |
| Eating and drinking places (not serving alcohol) | −1.21 (−2.19, −0.23) | −8.4 (−16.2, −0.7) | −6.8 (−12.4, −1.3) |
| Accommodations | −0.44 (−1.54, 0.67) | −1.3 (−9.9, 7.4) | −5.2 (−11.5, 1.0) |
| Medical care (at hospitals or clinics) | −0.56 (−1.46, 0.34) | −1.2 (−8.3, 5.8) | −6.4 (−11.6, −1.3) |
| Medical care (not at hospitals or clinics) | −0.33 (−1.29, 0.62) | −0.5 (−8.0, 7.1) | −4.9 (−10.4, 0.5) |
| Welfare | −0.27 (−1.18, 0.64) | −1.5 (−8.5, 5.6) | −4.5 (−9.7, 0.7) |
| Education and learning support | −0.71 (−1.60, 0.18) | −3.9 (−10.8, 3.1) | −6.0 (−11.1, −0.8) |
| Other services or public service | −0.66 (−1.54, 0.21) | −4.0 (−10.8, 2.8) | −6.3 (−11.3, −1.3) |
| **Employment type (reference: Regular employee)** | | | |
| Dispatched worker | 0.50 (0.07, 0.93) | 3.0 (−0.3, 6.3) | 1.5 (−0.8, 3.8) |
| Contract or entrusted employee | 0.22 (−0.03, 0.48) | 1.8 (−0.2, 3.9) | 0.9 (−0.4, 2.2) |
| Part-time or temporary worker | 0.08 (−0.10, 0.26) | −0.6 (−2.1, 0.9) | 0.3 (−0.6, 1.3) |
| **Company size, employees (reference: 1–4)** | | | |
| 5–99 | 0.41 (0.09, 0.73) | 3.1 (0.4, 5.7) | 2.2 (0.6, 3.9) |
| 100–999 | 0.55 (0.22, 0.87) | 4.5 (1.8, 7.2) | 2.4 (0.7, 4.0) |
| ≥1,000 | 0.33 (0.00, 0.66) | 2.2 (−0.5, 5.0) | 1.9 (0.2, 3.5) |
| Don't know | 0.52 (0.16, 0.88) | 4.5 (1.5, 7.5) | 1.9 (0.1, 3.8) |
| **Household type (reference: Living alone)** | | | |
| With spouse or partner, no child <18 | −0.38 (−0.57, −0.20) | −2.7 (−4.2, −1.2) | −1.2 (−2.2, −0.3) |
| With spouse or partner and child <18 | −0.72 (−0.91, −0.52) | −4.6 (−6.2, −3.0) | −2.9 (−4.0, −1.9) |
| Other | 0.62 (0.43, 0.82) | 4.2 (2.6, 5.7) | 2.8 (1.8, 3.9) |
| **Annual household income, million JPY (reference: <2)** | | | |
| 2 to <4 | −1.20 (−1.56, −0.85) | −7.6 (−10.2, −5.0) | −5.5 (−7.5, −3.4) |
| 4 to <6 | −1.98 (−2.33, −1.63) | −13.1 (−15.7, −10.6) | −8.4 (−10.4, −6.5) |
| 6 to <8 | −2.39 (−2.76, −2.03) | −16.6 (−19.3, −13.8) | −10.4 (−12.4, −8.4) |
| 8 to <10 | −2.62 (−3.00, −2.23) | −18.5 (−21.3, −15.6) | −10.8 (−12.9, −8.7) |
| ≥10 | −2.91 (−3.29, −2.53) | −20.9 (−23.8, −18.1) | −11.6 (−13.7, −9.5) |
| Prefer not to answer | −2.35 (−2.73, −1.96) | −16.1 (−19.0, −13.3) | −9.7 (−11.8, −7.6) |
| Don't know | −2.29 (−2.66, −1.91) | −15.9 (−18.7, −13.1) | −9.5 (−11.6, −7.4) |
| **Survey wave (reference: 2021)** | | | |
| 2022 | 0.10 (−0.15, 0.35) | −0.5 (−2.6, 1.7) | 1.9 (0.4, 3.3) |
| 2023 | 0.14 (−0.10, 0.39) | 0.1 (−1.9, 2.2) | 1.4 (0.0, 2.8) |
| 2024 | 0.74 (0.48, 1.00) | 4.7 (2.5, 6.9) | 4.5 (3.1, 6.0) |
| 2025 | 0.32 (0.13, 0.52) | 1.6 (−0.1, 3.3) | 2.2 (1.1, 3.3) |
| **Fixed at the first response** | | | |
| **Sex (reference: Men)** | | | |
| Women | 0.02 (−0.12, 0.16) | 0.8 (−0.3, 1.9) | −0.5 (−1.2, 0.2) |
| **Age group (2021 equivalent), years (reference: <20)** | | | |
| 20–29 | 0.59 (0.29, 0.90) | 3.9 (1.5, 6.3) | 2.1 (0.4, 3.9) |
| 30–39 | 0.41 (0.09, 0.74) | 2.2 (−0.4, 4.8) | 0.8 (−1.0, 2.7) |
| 40–49 | −0.11 (−0.43, 0.22) | −2.2 (−4.8, 0.4) | −0.6 (−2.4, 1.3) |
| 50–59 | −1.22 (−1.55, −0.89) | −10.6 (−13.3, −7.9) | −5.2 (−7.0, −3.4) |
| 60–69 | −3.07 (−3.41, −2.74) | −25.6 (−28.4, −22.8) | −11.3 (−13.1, −9.4) |
| ≥70 | −3.86 (−4.24, −3.49) | −32.8 (−36.1, −29.5) | −13.0 (−14.9, −11.1) |
| **Region (reference: Hokkaido)** | | | |
| Tohoku | 0.87 (0.51, 1.23) | 6.6 (3.8, 9.5) | 3.0 (1.1, 4.9) |
| Kanto | 0.30 (0.03, 0.58) | 2.8 (0.5, 5.1) | 0.8 (−0.6, 2.3) |
| Chubu | 0.51 (0.22, 0.80) | 5.0 (2.6, 7.4) | 1.5 (0.0, 3.0) |
| Kinki | 0.27 (−0.02, 0.55) | 2.5 (0.2, 4.9) | 0.7 (−0.8, 2.2) |
| Chugoku | 0.29 (−0.06, 0.64) | 3.7 (0.8, 6.6) | 0.4 (−1.4, 2.2) |
| Shikoku | 0.13 (−0.30, 0.56) | 1.5 (−2.0, 5.1) | 0.0 (−2.2, 2.3) |
| Kyushu/Okinawa | 0.36 (0.05, 0.68) | 3.5 (0.9, 6.2) | 0.7 (−0.9, 2.4) |
| **Education (reference: High school or less)** | | | |
| Vocational/junior college | −0.07 (−0.25, 0.11) | 0.1 (−1.4, 1.5) | −1.1 (−2.0, −0.2) |
| University or higher | −0.03 (−0.19, 0.13) | −0.1 (−1.4, 1.2) | −0.8 (−1.6, 0.1) |
| Other | 0.87 (0.04, 1.70) | 5.9 (−0.3, 12.2) | 2.8 (−1.6, 7.3) |

| Term | K6 score, points | K6 ≥5, percentage points | K6 ≥13, percentage points |
|---|---|---|---|
| Intercept | 6.72 (5.67, 7.78) | 51.6 (43.4, 59.9) | 23.1 (17.1, 29.0) |

Coefficients (95% CI) from within–between linear regression; 78,498 person-waves from 39,773 persons. 95% CIs are based on standard errors clustered by person.

Within-person terms are deviations of each indicator from the person's mean over the waves in the analysis; between-person terms are the person means. For K6 ≥5 and K6 ≥13, coefficients of linear probability models are multiplied by 100.

**Table S4. Proportion of person-waves in 2021–2024 with K6 observed at the next survey wave, by characteristics at the current wave**

| Characteristic at the current wave | Person-waves, n | K6 observed at the next wave, n (%) |
|---|---|---|
| All person-waves | 61,936 | 37,043 (59.8) |
| **Survey waves (current–next)** | | |
| 2021–2022 | 15,167 | 10,188 (67.2) |
| 2022–2023 | 16,259 | 9,572 (58.9) |
| 2023–2024 | 16,156 | 10,192 (63.1) |
| 2024–2025 | 14,354 | 7,091 (49.4) |
| **Frequency of working from home** | | |
| None | 48,488 | 29,032 (59.9) |
| Once a month | 1,832 | 1,055 (57.6) |
| 2–3 times a month | 2,082 | 1,197 (57.5) |
| Once a week | 2,313 | 1,365 (59.0) |
| 2–3 times a week | 3,457 | 2,110 (61.0) |
| 4–5 times a week | 2,085 | 1,303 (62.5) |
| Almost daily | 1,679 | 981 (58.4) |
| **K6 score category** | | |
| 0–4 | 36,998 | 22,574 (61.0) |
| 5–12 | 17,664 | 10,290 (58.3) |
| ≥13 | 7,274 | 4,179 (57.5) |
| **Sex** | | |
| Men | 33,399 | 20,659 (61.9) |
| Women | 28,537 | 16,384 (57.4) |
| **Age group (2021 equivalent), years** | | |
| <20 | 2,113 | 723 (34.2) |
| 20–29 | 14,391 | 7,132 (49.6) |
| 30–39 | 13,704 | 7,740 (56.5) |
| 40–49 | 13,644 | 8,863 (65.0) |
| 50–59 | 10,805 | 7,358 (68.1) |
| 60–69 | 5,849 | 4,198 (71.8) |
| ≥70 | 1,430 | 1,029 (72.0) |
| **Education** | | |
| High school or less | 14,068 | 8,385 (59.6) |
| Vocational/junior college | 12,707 | 7,534 (59.3) |
| University or higher | 34,816 | 20,938 (60.1) |
| Other | 345 | 186 (53.9) |
| **Employment type** | | |
| Regular employee | 40,286 | 24,221 (60.1) |
| Dispatched worker | 1,636 | 1,013 (61.9) |
| Contract or entrusted employee | 4,471 | 3,010 (67.3) |
| Part-time or temporary worker | 15,543 | 8,799 (56.6) |
| **Occupation** | | |
| Professional or technical | 14,524 | 8,731 (60.1) |
| Clerical | 16,668 | 10,408 (62.4) |
| Sales | 7,804 | 4,627 (59.3) |
| Service | 4,992 | 2,759 (55.3) |
| Security | 861 | 514 (59.7) |
| Production | 3,672 | 2,280 (62.1) |
| Transport or machine operation | 1,114 | 652 (58.5) |
| Construction or mining | 653 | 394 (60.3) |
| Carrying, cleaning or packaging | 1,904 | 1,166 (61.2) |
| Other | 9,744 | 5,512 (56.6) |
| **Company size, employees** | | |
| 1–4 | 2,108 | 1,298 (61.6) |
| 5–99 | 18,840 | 11,228 (59.6) |
| 100–999 | 16,373 | 9,965 (60.9) |
| ≥1,000 | 18,250 | 11,057 (60.6) |
| Don't know | 6,365 | 3,495 (54.9) |
| **Household type** | | |
| Living alone | 15,119 | 9,092 (60.1) |
| With spouse or partner, no child <18 | 17,307 | 11,348 (65.6) |
| With spouse or partner and child <18 | 15,298 | 8,485 (55.5) |
| Other | 14,212 | 8,118 (57.1) |
| **Annual household income, million JPY** | | |
| <2 | 3,296 | 1,939 (58.8) |
| 2 to <4 | 10,264 | 6,304 (61.4) |
| 4 to <6 | 12,646 | 7,784 (61.6) |
| 6 to <8 | 9,737 | 5,841 (60.0) |
| 8 to <10 | 6,928 | 4,172 (60.2) |
| ≥10 | 7,347 | 4,428 (60.3) |
| Prefer not to answer | 5,599 | 3,321 (59.3) |
| Don't know | 6,119 | 3,254 (53.2) |

All person-waves in the analytic sample in 2021–2024 are shown (61,936 person-waves from 31,038 persons). Those with K6 observed at the next survey wave formed the pairs used in the analysis (Table S5).

K6 at the next wave was counted as observed regardless of employment status at the next wave.

**Table S5. Characteristics at the current wave of the pairs used in the analysis of K6 at the next survey wave, by frequency of working from home at the current wave**

| Characteristic | Total | None | Once a month | 2–3 times a month | Once a week | 2–3 times a week | 4–5 times a week | Almost daily |
|---|---|---|---|---|---|---|---|---|
| Pairs, n | 37,043 | 29,032 | 1,055 | 1,197 | 1,365 | 2,110 | 1,303 | 981 |
| **Survey waves (current–next)** | | | | | | | | |
| 2021–2022 | 10,188 (27.5) | 7,721 (26.6) | 269 (25.5) | 373 (31.2) | 396 (29.0) | 685 (32.5) | 416 (31.9) | 328 (33.4) |
| 2022–2023 | 9,572 (25.8) | 7,468 (25.7) | 281 (26.6) | 296 (24.7) | 367 (26.9) | 539 (25.5) | 376 (28.9) | 245 (25.0) |
| 2023–2024 | 10,192 (27.5) | 8,224 (28.3) | 283 (26.8) | 298 (24.9) | 338 (24.8) | 505 (23.9) | 305 (23.4) | 239 (24.4) |
| 2024–2025 | 7,091 (19.1) | 5,619 (19.4) | 222 (21.0) | 230 (19.2) | 264 (19.3) | 381 (18.1) | 206 (15.8) | 169 (17.2) |
| **Sex** | | | | | | | | |
| Men | 20,659 (55.8) | 15,302 (52.7) | 735 (69.7) | 860 (71.8) | 952 (69.7) | 1,354 (64.2) | 839 (64.4) | 617 (62.9) |
| Women | 16,384 (44.2) | 13,730 (47.3) | 320 (30.3) | 337 (28.2) | 413 (30.3) | 756 (35.8) | 464 (35.6) | 364 (37.1) |
| **Age group (2021 equivalent), years** | | | | | | | | |
| <20 | 723 (2.0) | 638 (2.2) | 20 (1.9) | 21 (1.8) | 14 (1.0) | 13 (0.6) | 9 (0.7) | 8 (0.8) |
| 20–29 | 7,132 (19.3) | 5,379 (18.5) | 251 (23.8) | 279 (23.3) | 323 (23.7) | 416 (19.7) | 272 (20.9) | 212 (21.6) |
| 30–39 | 7,740 (20.9) | 5,842 (20.1) | 257 (24.4) | 294 (24.6) | 335 (24.5) | 480 (22.7) | 323 (24.8) | 209 (21.3) |
| 40–49 | 8,863 (23.9) | 6,896 (23.8) | 257 (24.4) | 300 (25.1) | 320 (23.4) | 521 (24.7) | 303 (23.3) | 266 (27.1) |
| 50–59 | 7,358 (19.9) | 5,838 (20.1) | 198 (18.8) | 210 (17.5) | 240 (17.6) | 421 (20.0) | 274 (21.0) | 177 (18.0) |
| 60–69 | 4,198 (11.3) | 3,539 (12.2) | 56 (5.3) | 74 (6.2) | 109 (8.0) | 224 (10.6) | 113 (8.7) | 83 (8.5) |
| ≥70 | 1,029 (2.8) | 900 (3.1) | 16 (1.5) | 19 (1.6) | 24 (1.8) | 35 (1.7) | 9 (0.7) | 26 (2.7) |
| **Education** | | | | | | | | |
| High school or less | 8,385 (22.6) | 7,539 (26.0) | 114 (10.8) | 139 (11.6) | 155 (11.4) | 184 (8.7) | 128 (9.8) | 126 (12.8) |
| Vocational/junior college | 7,534 (20.3) | 6,512 (22.4) | 148 (14.0) | 140 (11.7) | 151 (11.1) | 253 (12.0) | 158 (12.1) | 172 (17.5) |
| University or higher | 20,938 (56.5) | 14,843 (51.1) | 789 (74.8) | 915 (76.4) | 1,054 (77.2) | 1,660 (78.7) | 1,007 (77.3) | 670 (68.3) |
| Other | 186 (0.5) | 138 (0.5) | 4 (0.4) | 3 (0.3) | 5 (0.4) | 13 (0.6) | 10 (0.8) | 13 (1.3) |
| **Region** | | | | | | | | |
| Hokkaido | 1,583 (4.3) | 1,387 (4.8) | 32 (3.0) | 33 (2.8) | 40 (2.9) | 37 (1.8) | 27 (2.1) | 27 (2.8) |
| Tohoku | 2,269 (6.1) | 1,997 (6.9) | 37 (3.5) | 40 (3.3) | 44 (3.2) | 81 (3.8) | 34 (2.6) | 36 (3.7) |
| Kanto | 13,807 (37.3) | 9,401 (32.4) | 472 (44.7) | 567 (47.4) | 677 (49.6) | 1,216 (57.6) | 881 (67.6) | 593 (60.4) |
| Chubu | 6,208 (16.8) | 5,272 (18.2) | 165 (15.6) | 152 (12.7) | 179 (13.1) | 222 (10.5) | 108 (8.3) | 110 (11.2) |
| Kinki | 6,744 (18.2) | 5,341 (18.4) | 193 (18.3) | 236 (19.7) | 264 (19.3) | 379 (18.0) | 194 (14.9) | 137 (14.0) |
| Chugoku | 2,040 (5.5) | 1,802 (6.2) | 46 (4.4) | 57 (4.8) | 46 (3.4) | 54 (2.6) | 16 (1.2) | 19 (1.9) |
| Shikoku | 973 (2.6) | 897 (3.1) | 23 (2.2) | 15 (1.3) | 14 (1.0) | 14 (0.7) | 5 (0.4) | 5 (0.5) |
| Kyushu/Okinawa | 3,419 (9.2) | 2,935 (10.1) | 87 (8.2) | 97 (8.1) | 101 (7.4) | 107 (5.1) | 38 (2.9) | 54 (5.5) |
| **Employment type** | | | | | | | | |
| Regular employee | 24,221 (65.4) | 17,673 (60.9) | 892 (84.5) | 1,008 (84.2) | 1,116 (81.8) | 1,722 (81.6) | 1,047 (80.4) | 763 (77.8) |
| Dispatched worker | 1,013 (2.7) | 832 (2.9) | 23 (2.2) | 19 (1.6) | 20 (1.5) | 45 (2.1) | 47 (3.6) | 27 (2.8) |
| Contract or entrusted employee | 3,010 (8.1) | 2,439 (8.4) | 36 (3.4) | 60 (5.0) | 100 (7.3) | 166 (7.9) | 112 (8.6) | 97 (9.9) |
| Part-time or temporary worker | 8,799 (23.8) | 8,088 (27.9) | 104 (9.9) | 110 (9.2) | 129 (9.5) | 177 (8.4) | 97 (7.4) | 94 (9.6) |
| **Occupation** | | | | | | | | |
| Professional or technical | 8,731 (23.6) | 6,079 (20.9) | 334 (31.7) | 320 (26.7) | 380 (27.8) | 650 (30.8) | 557 (42.7) | 411 (41.9) |
| Clerical | 10,408 (28.1) | 7,641 (26.3) | 356 (33.7) | 409 (34.2) | 473 (34.7) | 820 (38.9) | 438 (33.6) | 271 (27.6) |
| Sales | 4,627 (12.5) | 3,410 (11.7) | 164 (15.5) | 218 (18.2) | 244 (17.9) | 331 (15.7) | 142 (10.9) | 118 (12.0) |
| Service | 2,759 (7.4) | 2,557 (8.8) | 28 (2.7) | 41 (3.4) | 50 (3.7) | 41 (1.9) | 17 (1.3) | 25 (2.5) |
| Security | 514 (1.4) | 480 (1.7) | 6 (0.6) | 6 (0.5) | 12 (0.9) | 7 (0.3) | 1 (0.1) | 2 (0.2) |
| Production | 2,280 (6.2) | 2,142 (7.4) | 26 (2.5) | 37 (3.1) | 30 (2.2) | 18 (0.9) | 11 (0.8) | 16 (1.6) |
| Transport or machine operation | 652 (1.8) | 601 (2.1) | 15 (1.4) | 10 (0.8) | 10 (0.7) | 10 (0.5) | 3 (0.2) | 3 (0.3) |
| Construction or mining | 394 (1.1) | 337 (1.2) | 5 (0.5) | 12 (1.0) | 18 (1.3) | 11 (0.5) | 6 (0.5) | 5 (0.5) |
| Carrying, cleaning or packaging | 1,166 (3.1) | 1,114 (3.8) | 9 (0.9) | 11 (0.9) | 12 (0.9) | 10 (0.5) | 5 (0.4) | 5 (0.5) |
| Other | 5,512 (14.9) | 4,671 (16.1) | 112 (10.6) | 133 (11.1) | 136 (10.0) | 212 (10.0) | 123 (9.4) | 125 (12.7) |

| Characteristic | Total | None | Once a month | 2–3 times a month | Once a week | 2–3 times a week | 4–5 times a week | Almost daily |
|---|---|---|---|---|---|---|---|---|
| **Company size, employees** | | | | | | | | |
| 1–4 | 1,298 (3.5) | 1,091 (3.8) | 14 (1.3) | 32 (2.7) | 27 (2.0) | 48 (2.3) | 32 (2.5) | 54 (5.5) |
| 5–99 | 11,228 (30.3) | 9,695 (33.4) | 195 (18.5) | 247 (20.6) | 260 (19.0) | 366 (17.3) | 234 (18.0) | 231 (23.5) |
| 100–999 | 9,965 (26.9) | 7,784 (26.8) | 280 (26.5) | 335 (28.0) | 430 (31.5) | 569 (27.0) | 345 (26.5) | 222 (22.6) |
| ≥1,000 | 11,057 (29.8) | 7,340 (25.3) | 526 (49.9) | 536 (44.8) | 583 (42.7) | 1,033 (49.0) | 632 (48.5) | 407 (41.5) |
| Don't know | 3,495 (9.4) | 3,122 (10.8) | 40 (3.8) | 47 (3.9) | 65 (4.8) | 94 (4.5) | 60 (4.6) | 67 (6.8) |
| Working ≥30 hours a week | 28,781 (77.7) | 21,815 (75.1) | 920 (87.2) | 1,035 (86.5) | 1,162 (85.1) | 1,842 (87.3) | 1,171 (89.9) | 836 (85.2) |
| Mainly desk work | 18,081 (48.8) | 11,887 (40.9) | 717 (68.0) | 834 (69.7) | 967 (70.8) | 1,715 (81.3) | 1,146 (88.0) | 815 (83.1) |
| **Household type** | | | | | | | | |
| Living alone | 9,092 (24.5) | 6,947 (23.9) | 271 (25.7) | 323 (27.0) | 349 (25.6) | 536 (25.4) | 380 (29.2) | 286 (29.2) |
| With spouse or partner, no child <18 | 11,348 (30.6) | 9,011 (31.0) | 287 (27.2) | 307 (25.6) | 402 (29.5) | 702 (33.3) | 364 (27.9) | 275 (28.0) |
| With spouse or partner and child <18 | 8,485 (22.9) | 6,222 (21.4) | 330 (31.3) | 393 (32.8) | 394 (28.9) | 556 (26.4) | 345 (26.5) | 245 (25.0) |
| Other | 8,118 (21.9) | 6,852 (23.6) | 167 (15.8) | 174 (14.5) | 220 (16.1) | 316 (15.0) | 214 (16.4) | 175 (17.8) |
| **Annual household income, million JPY** | | | | | | | | |
| <2 | 1,939 (5.2) | 1,727 (5.9) | 20 (1.9) | 35 (2.9) | 39 (2.9) | 57 (2.7) | 34 (2.6) | 27 (2.8) |
| 2 to <4 | 6,304 (17.0) | 5,425 (18.7) | 123 (11.7) | 129 (10.8) | 150 (11.0) | 198 (9.4) | 148 (11.4) | 131 (13.4) |
| 4 to <6 | 7,784 (21.0) | 6,188 (21.3) | 197 (18.7) | 269 (22.5) | 286 (21.0) | 412 (19.5) | 253 (19.4) | 179 (18.2) |
| 6 to <8 | 5,841 (15.8) | 4,473 (15.4) | 196 (18.6) | 223 (18.6) | 235 (17.2) | 352 (16.7) | 197 (15.1) | 165 (16.8) |
| 8 to <10 | 4,172 (11.3) | 3,041 (10.5) | 174 (16.5) | 176 (14.7) | 192 (14.1) | 291 (13.8) | 182 (14.0) | 116 (11.8) |
| ≥10 | 4,428 (12.0) | 2,711 (9.3) | 221 (20.9) | 250 (20.9) | 278 (20.4) | 497 (23.6) | 281 (21.6) | 190 (19.4) |
| Prefer not to answer | 3,321 (9.0) | 2,651 (9.1) | 66 (6.3) | 67 (5.6) | 110 (8.1) | 196 (9.3) | 131 (10.1) | 100 (10.2) |
| Don't know | 3,254 (8.8) | 2,816 (9.7) | 58 (5.5) | 48 (4.0) | 75 (5.5) | 107 (5.1) | 77 (5.9) | 73 (7.4) |
| K6 score, mean (SD) | 4.8 (5.7) | 4.7 (5.7) | 5.0 (5.4) | 5.9 (5.9) | 5.5 (5.6) | 4.8 (5.6) | 4.5 (5.5) | 4.7 (6.0) |
| **K6 score category** | | | | | | | | |
| 0–4 | 22,574 (60.9) | 17,949 (61.8) | 606 (57.4) | 617 (51.5) | 716 (52.5) | 1,257 (59.6) | 828 (63.5) | 601 (61.3) |
| 5–12 | 10,290 (27.8) | 7,911 (27.2) | 327 (31.0) | 382 (31.9) | 459 (33.6) | 611 (29.0) | 328 (25.2) | 272 (27.7) |
| ≥13 | 4,179 (11.3) | 3,172 (10.9) | 122 (11.6) | 198 (16.5) | 190 (13.9) | 242 (11.5) | 147 (11.3) | 108 (11.0) |

Values are n (column %) of pairs unless otherwise indicated; 37,043 pairs from 18,646 persons. A person can contribute up to four pairs.
Sex, age group, education and region were fixed at the first valid response in 2021–2025; age was converted to the 2021 equivalent.

**Table S6. Frequency of working from home by sex and by household circumstances at the first wave in the analysis**

| | Men | Women | Living with others, no child <18 | Living alone | Living with a child <18 |
|---|---|---|---|---|---|
| Persons, n | 20,058 | 19,715 | 19,202 | 10,138 | 10,433 |
| Person-waves, n | 41,765 | 36,733 | 38,529 | 19,540 | 20,429 |
| Persons observed in ≥2 waves, n | 10,268 | 8,788 | 9,343 | 4,645 | 5,068 |
| Changed WFH frequency category, n (% of persons in ≥2 waves) | 3,160 (30.8) | 1,719 (19.6) | 2,103 (22.5) | 1,342 (28.9) | 1,434 (28.3) |
| Reported different household circumstances later, n (% of persons in ≥2 waves) | – | – | 1,426 (15.3) | 876 (18.9) | 720 (14.2) |
| **Frequency of working from home, n (% of person-waves)** | | | | | |
| None | 30,956 (74.1) | 30,750 (83.7) | 31,356 (81.4) | 14,917 (76.3) | 15,433 (75.5) |
| Once a month | 1,594 (3.8) | 773 (2.1) | 944 (2.5) | 637 (3.3) | 786 (3.8) |
| 2–3 times a month | 1,798 (4.3) | 797 (2.2) | 1,026 (2.7) | 734 (3.8) | 835 (4.1) |
| Once a week | 1,970 (4.7) | 946 (2.6) | 1,273 (3.3) | 781 (4.0) | 862 (4.2) |
| 2–3 times a week | 2,660 (6.4) | 1,619 (4.4) | 1,886 (4.9) | 1,175 (6.0) | 1,218 (6.0) |
| 4–5 times a week | 1,551 (3.7) | 1,014 (2.8) | 1,093 (2.8) | 701 (3.6) | 771 (3.8) |
| Almost daily | 1,236 (3.0) | 834 (2.3) | 951 (2.5) | 595 (3.0) | 524 (2.6) |

Sex was fixed at the first valid response. Household circumstances were classified at the person's first wave in the analysis and applied to all of that person's waves.
WFH, working from home.

#### Supplementary Figures

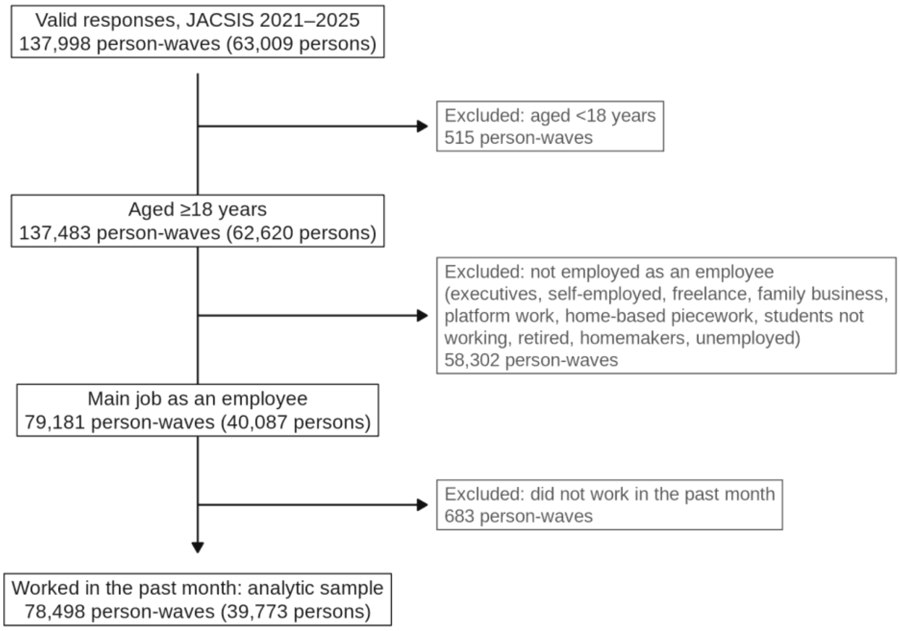


**Fig. S1** Selection of participants. Numbers are person-waves (numbers of persons in parentheses). No observations were excluded because of missing data on the frequency of working from home or on K6

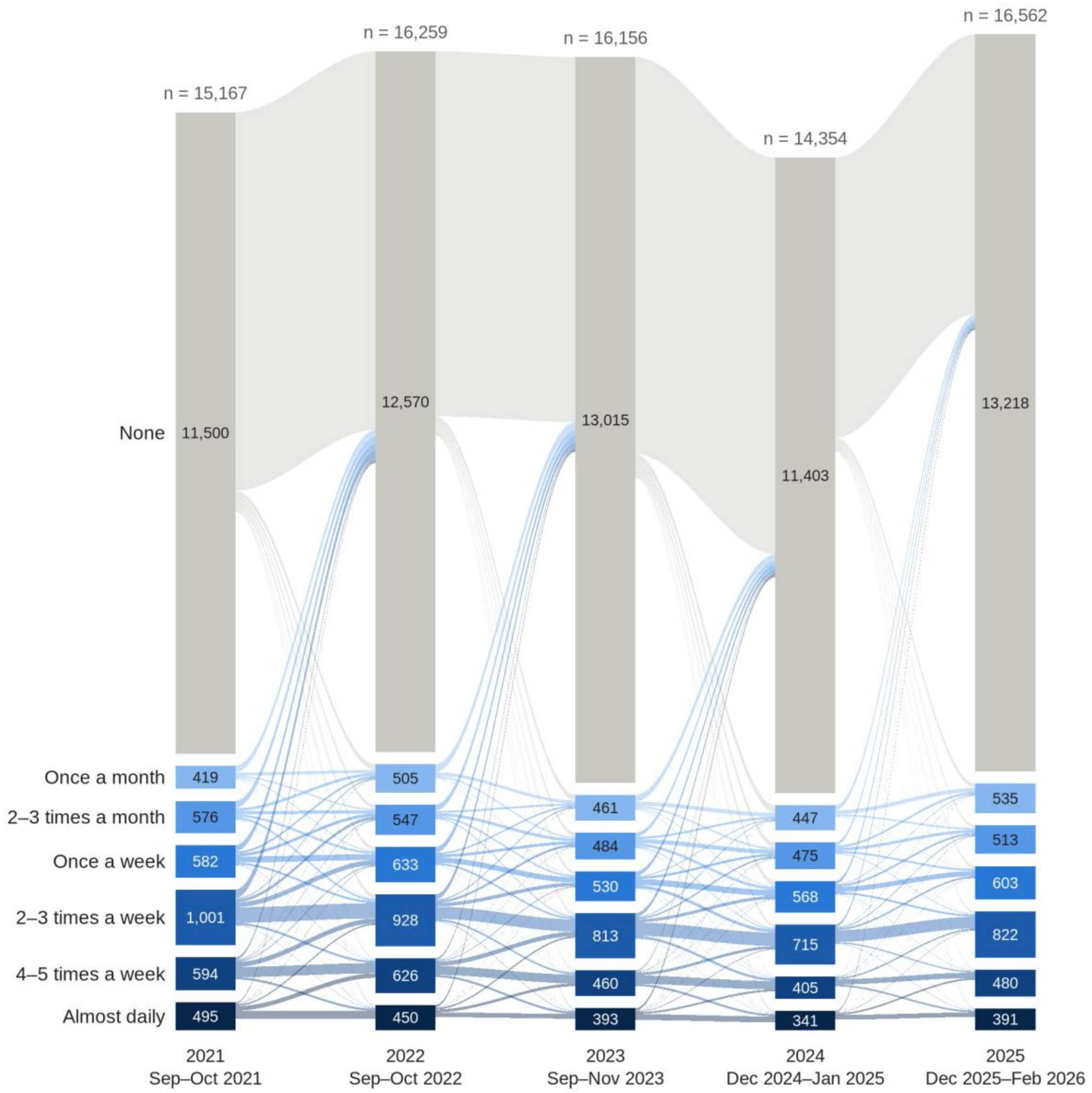


**Fig. S2** Frequency of working from home over the survey waves. The bar for each survey wave stacks all participants in the analytic sample in that wave by frequency of working from home, including those who took part only once and those who skipped a survey and took part again. Numbers in the bars are numbers of participants, and numbers above the bars are the total numbers of participants in each wave. The bands show transitions of the same persons who were in the analytic sample in both of two adjacent survey waves; the width of each band is proportional to the number of persons (34,168 pairs). Parts of a bar that are not connected to a band correspond to persons who were not in the analytic sample in the previous or the next wave (first participation, non-participation in the next survey, a change in employment status, or a skipped survey). 78,498 person-waves from 39,773 persons

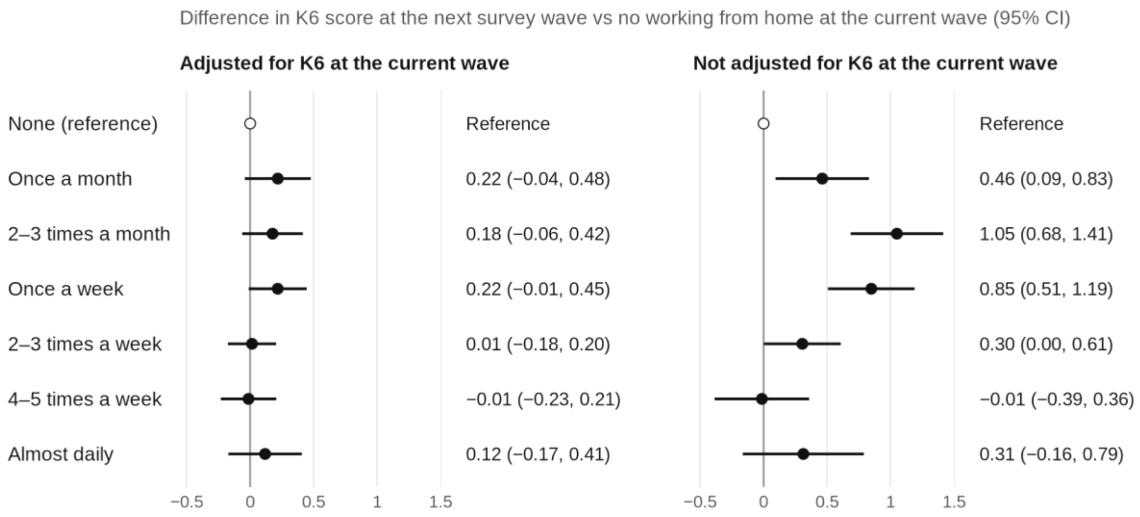


**Fig. S3** Frequency of working from home at the current wave and K6 at the next survey wave. Differences in the K6 score at the next survey wave compared with persons with no working from home at the current wave are shown as point estimates with 95% confidence intervals. Values on the right are point estimates (95% confidence intervals). The left panel shows the model adjusted for K6 at the current wave (natural spline), and the right panel shows the model without this adjustment; the horizontal axis is the same in both panels. Both models were adjusted for occupation, industry, employment type, company size, household type, and annual household income at the current wave, the pair of survey waves, and sex, age group, region, and education at the first response. Standard errors were clustered by person. 37,043 pairs from 18,646 persons

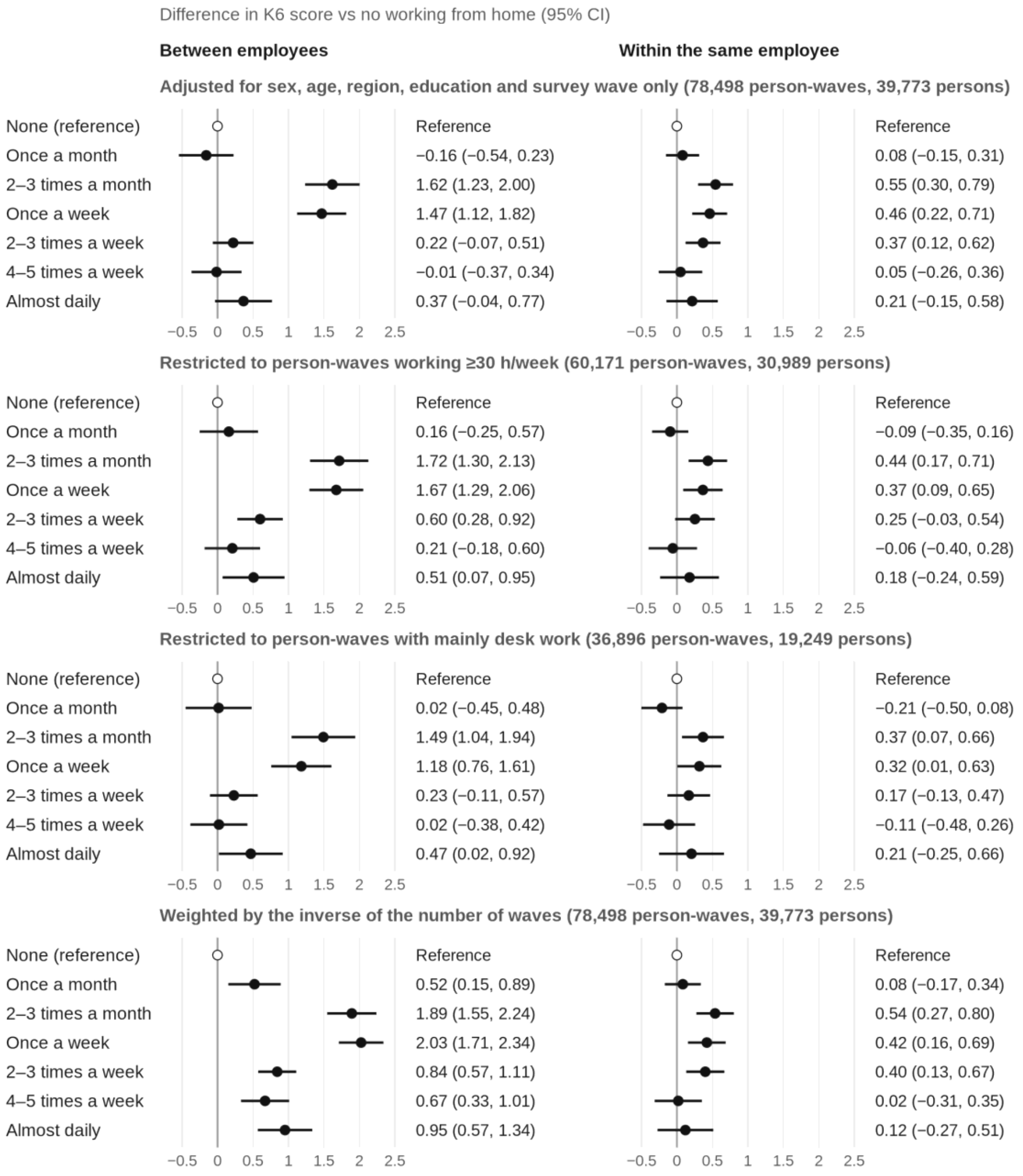


**Fig. S4** Supplementary analyses: between-person and within-person associations of the frequency of working from home with the K6 score. Differences in the K6 score compared with no working from home are shown as point estimates with 95% confidence intervals. Values on the right are point estimates (95% confidence intervals). From top to bottom, the panels show the analysis adjusted only for sex, age, region, education, and survey wave; the analysis restricted to person-waves working ≥30 hours a week; the analysis restricted to person-waves whose main work was desk work; and the analysis weighted by the inverse of the number of waves (so that the weights sum to the same total for each person). In the restricted analyses, person means were calculated from the person-waves remaining after the restriction. The horizontal axis is the same in all panels. Estimates from the main analysis are shown in Fig. 1 of the main text
